# Digital Biomarkers for Passive Remote Monitoring of Bipolar Disorder: Systematic Review

**DOI:** 10.1101/2025.09.23.25336470

**Authors:** Thomas P. Kutcher, Isha Chakraborty, Kristin Kostick-Quenet, Akane Sano, Nidal Moukaddam, Jeffrey A. Herron, Wayne K. Goodman, Sameer A. Sheth, Ashutosh Sabharwal, Nicole R. Provenza

**Author notes:** These authors contributed equally (Thomas P. Kutcher, Isha Chakraborty). Corresponding author: Nicole R. Provenza.

## Abstract

**Background:** Bipolar disorder (BD) features episodic shifts among (hypo)mania, depression, mixed states, and euthymia. Timely detection of mood transitions is difficult due to infrequent clinical touchpoints. Digital health technologies, including wearables and smartphones, offer a unique opportunity to passively and continuously monitor behavior and physiology that could reflect underlying mood dynamics in real-world settings.

**Objective:** We aim to systematically review passively collected digital biomarkers for BD mood states, characterize devices/modalities and analytic approaches, appraise risk of bias, and identify design gaps and priorities for clinical translation.

**Methods:** Following PRISMA guidelines (PROSPERO CRD42024607765), we searched MEDLINE, PsycINFO, Scopus, IEEE Xplore, and ACM Digital Library (February 7, 2025). We included peer-reviewed studies of adults with BD I/II that measured passively collected digital biomarkers and related them to depressive, (hypo)manic, mixed, or euthymic states. Active-only measures (e.g. lab tests, ecological-momentary assessment) and studies entangling BD with other diagnoses were excluded. Two independent reviewers screened studies and extracted study characteristics and results. We grouped digital biomarkers into categories and conducted narrative synthesis. Risk of bias was assessed with PROBAST (predictive models) and the Newcastle–Ottawa Scale (observational studies).

**Results:** Of 8,355 records, 45 studies met criteria. Most enrolled ≤50 participants (64%) and monitored ≤100 days (49%); 29% collected data only in-clinic. Nine biomarker domains emerged: physical activity, heart rate (HR), electrodermal activity (EDA), geolocation, keyboard use, light exposure, sleep, socialization, and speech. Consistent patterns linked depression to reduced mobility and social interaction, later/variable sleep, and lower daytime light; (hypo)mania was associated with higher and more variable activity, shorter/advanced sleep, and increased communication. Circadian features derived from sleep/activity repeatedly aided prediction. EDA tended to be lower in depression; HRV findings were mixed across settings and methods. Keyboard and speech features (e.g., timing, prosody) showed associations and performed well in classifiers. Fifteen studies used ML; several reported strong performance for episode prediction/classification (AUROC ≈0.80–0.98 in larger cohorts), yet external validation was absent, samples were small, monitoring windows were often short relative to episode timescales, clinical labels were infrequent/misaligned, and missingness was rarely modeled despite likely informativeness.

**Conclusions:** Passive digital biomarkers for BD show promise, with the most robust signals aligning with DSM-5 behavioral and circadian features (sleep–wake patterns, activity/mobility, socialization/geolocation, and speech). To move from promise to practice, future studies should adopt longer within-subject monitoring, align label cadence with sensing granularity, standardize features/reporting, pre-register analyses, externally validate models, minimize data to protect privacy, and expand physiological measurement beyond heart rate and electrodermal activity. These steps are essential to develop reliable, actionable tools for earlier detection and management of BD mood episodes.

## Introduction

Bipolar disorder (BD) is a recurrent chronic affective disorder marked by wide fluctuations in mood, energy, and activity that affects more than 1% of the world population [1]. BD often results in significant functional impairment and poor quality of life, imposing a significant burden on caregivers and society at large [1]. Patients chronically fluctuate between mood states, including depression, mania and/or hypomania, mixed states (e.g., irritable mood and elevated energy), and euthymia (stable mood) [1]. Real-life mood fluctuations in BD can follow various patterns, forming the basis for the concept of bipolar spectrum disorder. Based on episode severity and duration, mood episodes are classified as manic, depressive, or hypomanic, while intermediate patterns remain without named subtypes. Manic episodes are marked by hyperactivity, pressured speech, decreased need for sleep, expansive mood, risky behavior, and sometimes psychotic symptoms [2]. Hypomania is a milder form of mania in which functioning is less impaired [2]. Depressive episodes are marked by decreased energy, sadness, social withdrawal, insomnia or hypersomnia, and low self-esteem [2]. The DSM-5 subdivides BD into BD types I and II (BDI, BDII). BDI consists of manic and depressive episodes, and BDII consists of hypomanic and depressive episodes. Across both subtypes, patients experience euthymia about half of the time. In BDI, patients experience depression 32% and mania/mixed states 14.8% of the time, respectively. In BDII, patients experience depression 50% and hypomania/mixed states 3.6% of the time, respectively [3,4].

BD mood state fluctuations can threaten personal well-being. BD is marked by a suicide rate 20 times higher than that of the general population [5], and suicidality most commonly occurs in depressive and mixed states [6]. Additionally, mania and hypomania may lead to impairments in psycho-social function, reckless behavior (e.g. excessive spending, promiscuity, harming others), psychosis, increased risk of depression relapse, and increased caregiver burden [1]. Generally, clinicians become aware of patient mood state fluctuations through direct patient contact via clinic visits or phone calls [7]. Mood fluctuations in BD are extremely difficult to treat as episodes largely occur outside of clinical observation [8]. If the clinician is made aware of acute depression and (hypo)mania, the patient can then be treated with the appropriate intervention (e.g. pharmacological, behavioral) [2]. Taken together, there is an urgent need for new methods to remotely monitor the mood states of BD patients outside of the clinic to enable clinical intervention during (or even before) a relapse or episode recurrence.

Recently, studies have investigated the potential of using remote monitoring tools to facilitate BD treatment. Thus far, studies leveraging these tools have not yet demonstrated any significant improvement in patient outcomes [9,10]. Self-monitoring approaches have historically been limited by both poor patient insight and the high level of patient engagement required for data collection, leading to lapses in compliance [11]. Although specific diagnostic criteria of mania and depression (e.g., decreased need for sleep in mania; insomnia or hypersomnia in depression) are potentially measurable via wearable devices, evidence reliably linking these remotely-collected measures to specific mood states is inconclusive. While remote passive monitoring methods have demonstrated some preliminary promise, a greater understanding of objectively measurable behavioral or physiological signals (digital biomarkers) of BD mood states is necessary to enhance insights beyond what is learned from patient self-reports [7].

In the past five years, several reviews have been published that are relevant to remote monitoring in BD. A 2021 review explored the state of using portable technologies to monitor BD patients, focusing mainly on methodological approaches and less so on results [12]. Another 2021 review focused on the specific use of smartphone apps to remotely monitor BD symptoms, looking at both active and passive monitoring practices [13]. However, to our knowledge, no reviews have focused on discussing digital biomarkers of BD mood states that can be remotely and passively monitored, nor included discussion of the many recent studies that have implemented emerging machine learning (ML) and artificial intelligence (AI) based approaches that offer promising potential for monitoring mood states.

Our objective is thus to systematically review the literature to better understand the state of digital biomarkers that can be used for remote passive monitoring of BD. An improved understanding of the existing literature will lead to a better-informed implementation of remote monitoring practices in psychiatry and identify future work required to validate digital biomarkers of BD, thus improving patient outcomes.

## Methods

This review follows the Preferred Reporting Items for Systematic Reviews and Meta-Analyses (PRISMA) guidelines, as reported in Supplementary Table 4 [14]. All methods were pre-registered in the Prospective Register of Systematic Reviews (PROSPERO ID: CRD42024607765).

### Inclusion and Exclusion Criteria

Included articles were published, peer-reviewed studies that focused on adults with BD (BDI and BDII), established digital biomarkers (defined by the FDA as a characteristic or set of characteristics collected from digital health technologies that is measured as an indicator of normal biological processes, pathogenic processes, or responses to an exposure or intervention) [15] of (hypo)manic, depressive, mixed, or euthymic mood states in BD, and measured digital biomarkers passively (e.g. using devices such as wearables that are capable of passively collecting sleep and activity data without explicitly needing patient input). Studies that established digital biomarkers solely through active monitoring (patient self-reports, lab tests) or entangled results of a BD cohort with another group (major depressive disorder, schizophrenia, etc.) were excluded.

### Search Strategy

Five electronic databases were searched (MEDLINE, PsycINFO, Scopus, IEEE Xplore, and ACM Digital Library) on November 1, 2024 and updated on February 7, 2025. Search terms included a combination of keywords related to (1) population, (2) digital biomarkers, and (3) monitoring. The search was limited to after 2000 as remote monitoring technologies (e.g. smartphones, wearables) were not widely available before then. All search terms are detailed in the Supplementary Methods.

### Study Selection

Article screening was carried out by independent researchers T.K. and I.C. using the online tool Rayyan [16]. The first step was deduplication, where Rayyan assisted with flagging included studies with similar titles, abstracts, and authors for removal. The second step was title and abstract screening, where any titles or abstracts in misalignment with the inclusion criteria were excluded. For the third step, full-texts were extracted and evaluated against the inclusion criteria. Authors T.K. and I.C. independently carried out all steps of the study selection process and resolved conflicts through discussion when necessary.

### Data Extraction and Analysis

The following variables were extracted from all included studies where available: study identification, main findings, study design, methods (device/app, measurements, data characteristics), analytic approach (predictive [ML/AI] vs. associative [traditional inference]), clinical scales and their sampling frequency, and sample characteristics. For predictive studies, we reported the authors primary performance metrics (e.g., AUROC, accuracy, sensitivity/specificity, F1-score), and for associative studies, effect-size estimates (e.g., β, odds ratios, e^B^) with confidence intervals or p-values exactly as provided by authors. Where multiple metrics were given, we prioritized AUROC for classification tasks and standardized effect sizes for associations. Subsequently, included studies were assigned digital biomarker categories (e.g., speech, geolocation, etc.) and device categories (e.g., smartphones, wearables, etc.). To convey results, a narrative synthesis approach was used, focusing on thematic analysis of digital biomarker categories, device types, study outcomes, and methodology, while considering study design heterogeneity and potential biases. Additionally, authors T.K. and I.C. independently assessed the risk of bias in all studies. For studies developing or evaluating predictive models, we applied the Prediction Model Risk Of Bias Assessment Tool (PROBAST) [17] which evaluates issues related to study participants, predictors, outcome definitions, and analytic procedures. For non-predictive observational studies, we used the Newcastle-Ottawa Scale [18] assessing methodological quality based on participant selection, comparability of groups, and reliability of outcomes measurement. Potential biases and limitations regarding evidence certainty were qualitatively discussed.

## Results

### Search Results

The search identified 8355 articles, which were screened as detailed in Figure 1. Overall, 45 studies were included in the review.

**Figure 1:**
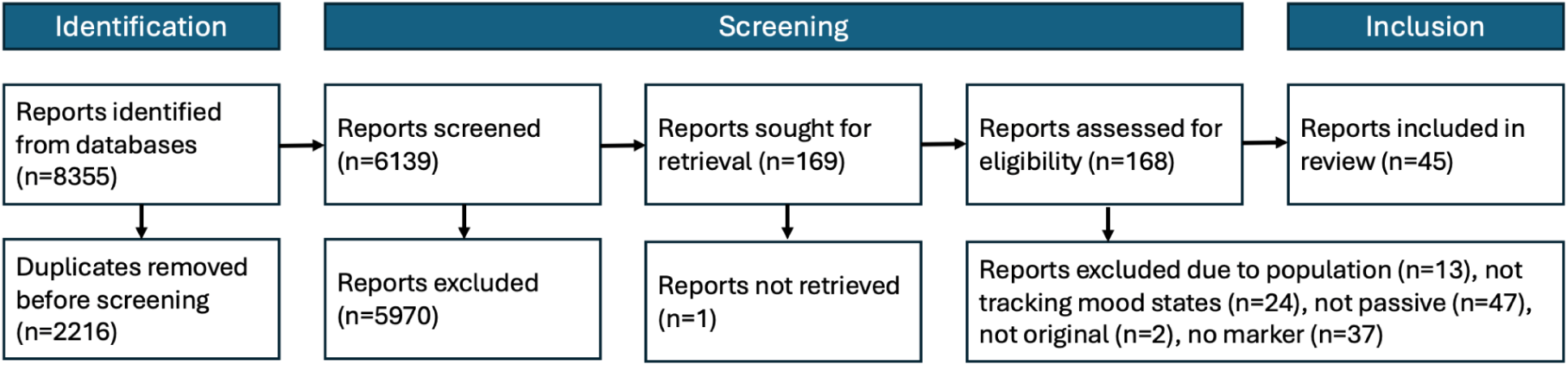
PRISMA study selection flowchart. This flowchart shows the number of articles involved in every step of the screening process.

### Study Characteristics

Table 1 summarizes the characteristics of the 45 studies included in this review, further detailed in Supplementary Table 1. Study sample sizes ranged from 6 [19] to 274 [20] participants, and most studies (64%) enrolled 50 or fewer participants. Study designs were heterogeneous, especially in study duration, setting, analysis methods, and clinical scales. 22 studies (49%) monitored patients for 100 days or less. 13 studies (29%) performed sensor measurements exclusively in a clinical setting (e.g., recording conversations during in-clinic evaluations). Some studies relied on inter-, rather than intra-, patient comparisons. Five studies (11%) captured only acute episodes rather than mood state changes within the same patient (e.g., the transition from mania to euthymia). Most studies (42 studies, 93%) used smartphones or wearable devices to perform measurements. Nearly all wearable devices used were commercial products, with the exception of three studies from a group that employed a custom ECG setup [21]. Analytical approaches varied substantially across studies; 15 studies (33%) utilized machine learning (ML) and artificial intelligence (AI)-based methods (e.g., supervised classifiers or predictive algorithms), while the remaining studies employed traditional statistical inference techniques (e.g., correlation analyses, regression modeling, or hypothesis-driven statistical tests) to examine associations between digital biomarkers and mood states. Finally, there was also heterogeneity in clinical scales used to measure mood states. For symptoms of mania or hypomania, 35 of the 40 studies used the Young Mania Rating Scale (YMRS). The remaining five studies assessed these symptoms using either the Bech-Rafaelsen Mania Scale (BRMS; 3 studies) or the Altman Self-Rating Mania Scale (ASRM; 2 studies). Depressive symptoms were assessed using the Hamilton Depression Rating Scale (HDRS) or Montgomery-Åsberg Depression Rating Scale (MADRS) in 33 studies, while the remaining seven studies utilized alternative instruments such as the Quick Inventory of Depressive Symptomatology (QIDS), Inventory of Depressive Symptomatology–Clinician Rated (IDS-C), Patient Health Questionnaire-8 (PHQ-8), or Patient Health Questionnaire-9 (PHQ-9). Scales were employed at varying frequencies and were subsequently associated with digital biomarkers.

**Table 1:** Study characteristics.

| *Category* | *Number of studies (%)* |
| --- | --- |
| **Publication year** |  |
| 2014 - 2016 | 12 (27%) |
| 2017 - 2019 | 11 (24%) |
| 2020 - 2022 | 9 (20%) |
| 2023 - 2025 | 13 (29%) |
| **Sample size** |  |
| 1 - 20 | 15 (33%) |
| 21 - 50 | 14 (31%) |
| 51 - 100 | 5 (11%) |
| 101 - 200 | 9 (20%) |
| 200 + | 2 (4%) |
| **Study duration (days)** |  |
| 1 | 1 (2%) |
| 2 - 7 | 10 (22%) |
| 8 - 30 | 3 (7%) |
| 31 - 100 | 9 (20%) |
| 101 - 365 | 13 (29%) |
| 366 + | 4 (9%) |
| Not reported | 4 (9%) |
| **Study setting** |  |
| Out of clinic | 32 (71%) |
| In clinic | 13 (29%) |
| **Measure intra-individual state change** |  |
| Yes | 39 (87%) |
| No | 6 (13%) |
| **Sensing device** |  |
| Wearable | 22 (49%) |
| Smartphone | 20 (44%) |
| Photometer | 2 (4%) |
| Microphone | 1 (2%) |
| **Types of wearable devices** |  |
| Empatica E4 | 5 (23%) |
| Fitbit | 4 (18%) |
| Actiwatch | 7 (32%) |
| Custom t-shirt | 3 (14%) |
| Polar RS | 1 (5%) |
| Oura Ring | 1 (5%) |
| Actiheart | 1 (5%) |
| **Predictive (ML/AI) or Associative**<br>**(traditional inference)** |  |
| Predictive | 15 (33%) |
| Associative | 30 (67%) |

### Digital Biomarkers Cluster Into 9 Distinct Categories

Overall, we divided the digital biomarkers of mood states detailed by the studies included in this review into nine distinct categories, as detailed in Table 2, according to standard methods used for measurement. Below, we discuss each of the categories, focusing on the methods used to define the digital biomarkers and the significance of the digital biomarkers as established through significant correlations with mood states or importance in classification algorithms. For studies that used multiple features in classification algorithms, we discuss the most important features for accurate mood state classification in this section and elaborate on the methods in the *Multivariable Modeling* section.

**Table 2:** Categories of digital biomarkers of bipolar disorder mood states.

| Digital Biomarker Category | Num. of studies (%) | Measurement Methods | Example Digital Biomarkers |
| --- | --- | --- | --- |
| Electrodermal Activity | 3 (7%) | Wearable sensors | Mean electrodermal activity (EDA), EDA peaks per minute, and EDA peaks mean amplitude |
| Geolocation | 6 (13%) | Smartphone GPS, cell tower, or WiFi | Change in smartphone cell tower ID, change in smartphone WiFi, global positioning system (GPS) |
| Heart Rate | 10 (22%) | Wearable sensors | Mean heart rate, heart rate variability (HRV), root mean square of HRV |
| Keyboard Usage | 4 (9%) | Smartphone keyboard app | Backspace rate, typing speed, accelerometer displacement, number of typing sessions, autocorrect rate |
| Light Exposure | 6 (13%) | Smartphone (or external) photometer | Average daytime or nighttime illuminance (lux) |
| Physical Activity | 12 (27%) | Wearable sensors | Acceleration, energy expenditure, metabolic equivalents (METs) |
| Sleep | 7 (16%) | Wearable sensors | Total sleep time, sleep efficiency, sleep variability |
| Socialization | 5 (11%) | Smartphone | Number of incoming and outgoing calls and texts, call duration, number of unique contacts |
| Speech | 9 (20%) | Smartphone (or external) microphone | Speech latencies, pitch, variance, amplitude |

#### Electrodermal Activity

Electrodermal activity (EDA) reflects autonomic nervous system function via skin conductance through sweat gland activity. Three studies used wearable devices to establish EDA digital biomarkers. Anmella et al. (2024) reported lower mean EDA levels and fewer EDA peaks per minute during bipolar depressive states compared to mania and euthymia, with metrics returning to higher levels post-clinical remission (all comparisons statistically significant: ANOVA, p≤.004). Also, Anmella et al. (2023) and Côté Allard et al. (2022) identified reduced mean EDA and peak frequency (captured by the Empatica E4 wearable) as important digital biomarkers for bipolar depression state classification in an acute clinical setting [22,23]. These findings suggest that EDA may serve as a relevant physiological digital biomarker, particularly sensitive for depressive episodes.

#### Geolocation

Geolocation, analyzed in six studies, was measured using changes in smartphone GPS, cell tower, or WiFi. Faurholt-Jepsen et al. (2014) and Faurholt-Jepsen et al. (2016a) found that depressive symptoms (HDRS scores) were associated with fewer cell tower transitions (*β*=-0.56, p=0.004) while manic symptoms (YMRS scores) were associated with increased cell tower transitions (*β*=0.4, p=0.04) [24,25]. Extending this finding, Faurholt-Jepsen et al. (2021) determined that depressive patients moved significantly less per day (*β*=-0.27, p=0.006), spent more time at individual stops (*β*=79.62, p<0.001), and had less location entropy (*β*=-0.083, p<0.001) compared to euthymia [26]. Palmius et al. (2017) used ten geolocation-based digital biomarkers across varying timescales to predict QIDS scores using linear regression (Mean Absolute Error (MAE)=3.73, *p*<10^-4^) and classified depressive episodes with a median F1 score of 0.857 [27]. Beiwinkel et al., (2016) found that reduced mobility (inferred by fewer smartphone cell tower movements) correlated with both depression (*β*=-0.11, p=0.03) and mania (*β*=-0.17, p<0.001) in 13 patients outside the clinic [28]. In summary, these studies indicate that a reduction in geographic movements may be a consistent digital biomarker of bipolar depression. However, the evidence linking these digital biomarkers with mania is mixed.

#### Heart Rate Variability (HRV)

Heart rate (HR) and heart rate variability (HRV) were analyzed in 11 studies using wearable sensors. Faurholt-Jepsen et al. (2017) reported significantly higher HRV during manic states compared to depressive (18% increase; e^B^=1.18, p<0.001) and euthymic states (17% increase; e^B^=1.17, p<0.001) [29]. Contrastingly, Wazen et al. (2018) observed decreased HRV in mania relative to euthymia (Cohen’s d=0.668) [30]. These contradictory findings may be attributed to significant differences in methodologies: Faurholt-Jepsen et al. (2017) monitored 16 outpatients using the Actiheart wearable over three consecutive days [29], while Wazen et al. (2018) monitored 19 inpatients using the Polar RS 800 frequency meter over 20-minute periods [30]. Corponi et al. (2024) found strong evidence for positive changes in HRV as a marker of symptom resolution (95.175% probability of positive direction) regardless of depression or (hypo)mania [31].

Three inpatient studies employed a custom wearable shirt with integrated electrodes. Valenza et al. (2014) demonstrated the feasibility of HRV-based mood state recognition by developing personalized Markov chain models from HRV data collected over 18 hours with a mood state classification accuracy of up to 95.8% [21]. Building on this work, Valenza et al. (2016) developed support vector machine learning models for next-day mood forecasting (euthymia/non-euthymia), achieving prediction accuracies as high as 83.3% (69% on average) [32]. Nardelli et al. (2017) observed reduced HR pattern complexity (p<0.05) as measured by the entropy of the R-R interval series in both depressive and hypomanic states [33].

Five studies found that HR based digital biomarkers were important features in mood state classification algorithms [22,23,34–36]. Notably, Lipschitz et al. (2024) found heart rate to be the most important predictor of (hypo)mania (AUROC=0.852) and among the top predictors of depression (AUROC=0.860) using the Fitbit Inspire in real-world contexts [36]. Overall, HR and HRV show promise as digital biomarkers for BD mood states relative to euthymia, though findings on HRV vary by methodology.

#### Keyboard usage

Four studies used the “BiAffect” app on Android smartphones to passively capture keyboard metrics. Liu et al. (2024) used a 3-class model of backspace rates (Low, Medium, High). They found that the Medium group compared with the Low group had significantly higher ratings of depression (*β*=2.32, p=0.008), and the High group compared with the Low group was associated with both nonzero ratings (*β*=1.91, p=0.02) and higher ratings of mania (*β*=1.46, p<0.001) [37]. Stange et al. (2018) found that instability of typing speed (root mean square successive difference of each day’s average typing speed) predicted elevated future symptoms of depression (R^2^=0.70, p=0.03) but not mania [38]. Zulueta et al. (2018) expanded on this work by examining other keyboard metrics. Using a linear mixed-effects model, depression symptoms correlated with greater inter-key delay (*β*=2.88, p=0.003), session count (*β*=2.18, p=0.003), autocorrect rate (*β*=2.67, p=0.004), and accelerometer displacement while typing (*β*=3.20, p=0.002) [39]. Using an ordinary least squares model, mania symptoms were correlated with decreased backspace rates (*β*=-0.30, p=0.014) and accelerometer displacement while typing (*β*=0.39, p=0.003) [39]. Utilizing the same “BiAffect” typing dataset, Cao et al. (2017) applied a deep learning architecture utilizing gated recurrent units to classify bipolar depression with 90% accuracy [40]. Overall, various keyboard metrics, including typing variability, backspace use, and accelerometer displacement, are associated with bipolar depressive and manic episodes, demonstrating potential for digital monitoring and symptom classification.

#### Light exposure

Six studies investigated the relationship between light exposure patterns and mood episodes. Four of these studies were part of the APPLE (Association between the Pathology of Bipolar Disorder and Light Exposure in Daily Life) cohort based in Japan [41]. In these studies, over 100 BD patients were monitored for seven days with photometers in the bedroom and wearables (Actigraph), and were then followed up at one and two years [41]. Esaki et al. (2020) and Esaki et al. (2022) found that nighttime bedroom light exposure was significantly associated with current and future manic episodes [41,42]. Specifically, Esaki et al. (2020) found that patients with an average light intensity at night exposure of ≥3 lux had a higher prevalence of a hypomanic state (Odds Ratio (OR): 2.15, p=0.02) [41], while Esaki et al. (2022) used a Cox proportional-hazards model to find that the probability for manic/hypomanic episode relapse was significantly higher when the average nighttime illuminance was ≥3 lux (Hazards Ratio=2.54, 95% confidence interval [CI]=1.33, 4.84) [42].

Additionally, Esaki et al. (2019) and Esaki et al. (2021a) found that higher daytime light intensity was linked to lower depression severity and a lower likelihood of depression relapse [43,44]. Specifically, Esaki et al. (2019) found that the highest tertile group in average daytime light intensity has a significantly lower odds ratio for a depressed state than the lowest tertile group (OR=0.33, p=0.009) [43]. Esaki et al. (2021a) found that a higher average illuminance and longer time >1000 lux during daytime exhibited a significant decrease in relapse into depressive episodes (per log lux and per log min; Hazards Ratio=0.65 and 0.61; 95% CI=0.49, 0.86 and 0.47, 0.78, respectively) [44].

Two studies from the Mood Disorder Cohort Research Consortium (MDCRC) [45] based in Korea supported prior results by determining light exposure during nighttime and daytime as measured by smartphone photometers to be one of the most important digital biomarkers for classifying both manic and depressive episodes [34,35]. As such, deviations in light exposure patterns may offer an indication of both current and future (hypo)manic and depressive mood states.

#### Physical Activity

Twelve studies used wearable sensors to measure physical activity levels through accelerometry. Ebner-Priemer et al. (2020) found that daily fluctuations in mood were best predicted by changes in same-day activity. Greater activity levels correlated with manic symptoms (*β*=0.123, CI=0.075, 0.170) while depressive symptoms correlated with less activity (*β*=-0.152, CI=-0.190, 0.113) as measured by smartphone physical activity measures including GPS travel distances, number of steps, and velocity of movement [46]. Faurholt-Jepsen et al. (2016b) reported that manic and mixed states have higher energy expenditure (kJ/kg/day, *β*=14.8, p=0.003) compared to euthymia, and higher energy expenditure (kJ/kg/day, *β*=16.6, p=0.020) and trunk acceleration (m/s^2^, *β*=-0.064, p=0.030) compared to depression [47]. Likewise, Gershon et al. (2016) found that depressive states were characterized by reduced overall activity (p<0.05) and a later activity onset with a midday activity spike and low evening activity (p<0.05) [48]. Esaki et al. (2021b) demonstrated that later daily peaks in physical activity rhythms (onset of the 10 consecutive hours with the highest accelerometry amplitude) were significantly associated with increased risk of future depressive episode relapses (Hazard Ratio=1.11, CI=1.00, 1.22) [49]. Additionally, these studies consistently demonstrated that manic episodes were marked by greater overall and more variable physical activity. In particular, Jakobsen et al. (2022) showed that manic episodes had greater variability (as defined by sample entropy) in motor activity within-subject compared to euthymia (p=0.025) [50]. Ortiz et al. (2025) found that within-day activity variability achieved high performance in predicting hypomanic episodes with a median time before onset of 2.5 days (balanced accuracy=0.89±0.11) [51].

Five studies found that physical activity-related digital biomarkers were important predictors in multivariable mood state classification algorithms. Two studies from the Mood Disorder Cohort Research Consortium (MDCRC) [45] determined steps at different times of the day, particularly during nighttime, as one of the most important digital biomarkers for classifying both manic and depressive episodes [34,35]. Additionally, Lipschitz et al. (2024) found that minutes spent “very active” per day was an important (top 7/17) marker for classification of both depression (AUROC=0.860) and (hypo)mania (AUROC=0.852), while minutes spent sedentary was important only for classification of depression [36]. In studies of acute inpatients wearing Empatica E4 wearable devices, both Cote-Allard et al. (2022) and Anmella et al. (2023) found acceleration to be one of the most important digital biomarkers in distinguishing mood episodes [22,23]. In summary, physical activity levels consistently correlate with bipolar mood states: manic episodes are characterized by higher and more variable activity, while depressive episodes show reduced activity and altered timing.

#### Sleep

Seven studies used wearable sensors to establish sleep digital biomarkers of BD mood states. Ebner-Priemer et al. (2020) demonstrated that total sleep duration and wakeup time were negatively associated with same-day manic symptoms (*β*=-0.098, CI=-0.157, −0.040) [46]. In the APPLE cohort, Esaki et al. (2023) found that variability in total sleep time was strongly associated with future increased mood episode relapse rates (Hazards Ratio=1.407, CI=1.057, 1.873 [52], and Esaki et al. (2024) found a significant association between daytime napping and depressive symptoms (number of nap days: OR=3.66, CI=1.32, 1.17; nap duration: OR=3.14, CI=1.12, 8.81) [53]. Ortiz et al. (2025) found that within-night sleep variability achieved high performance in predicting hypomanic episodes (balanced accuracy = 0.87 ± 0.13) and was the earliest indicator of an oncoming hypomanic episode with a median time before onset of 3 days [51].

Three studies from the Mood Disorder Cohort Research Consortium (MDCRC) [45] used the Fitbit Charge to measure sleep-related digital biomarkers in over 100 BD patients outside the clinic. Lim et al. (2024) used 36 digital biomarkers, all based on sleep and sleep-based circadian rhythms, and Extreme Gradient Boosting (XGBoost) to predict next-day mood episodes in 111 BD patients (AUROC=0.80, 0.98, and 0.95, for depressive, manic, and hypomanic episodes, respectively) [54]. Their study found that circadian phase shifts, as estimated using the sleep/wake time series, were the most significant predictors across depressive, manic, and hypomanic episodes, with delays associated with depressive episodes and advances associated with manic episodes [54]. Cho et al. (2019) and Lee et al. (2023) determined sleep quality, length, onset deviations, and offset deviations to be important in mania and depression classification [34,35]. These studies emphasize the predictive utility of sleep patterns and circadian rhythm disruptions (with shorter sleep and advanced rhythms linked to mania, and longer, variable sleep and delayed rhythms linked to depression), in anticipating mood transitions.

#### Socialization

Five studies linked smartphone-based socialization behaviors and mood symptoms in BD. Beiwinkel et al. (2016) found that fewer outgoing texts were predictive of more severe depressive symptoms (*β*=-0.28, p<0.001), while increased outgoing texts were associated with increased manic symptoms (*β*=0.48, p=0.03) [28]. Faurholt-Jepsen et al. (2015b) observed that a longer duration of incoming calls (seconds/day: *β*=17.15, p=0.037) and a longer duration of outgoing calls (seconds/day: *β*=26.33, p=0.006) was associated with depressive symptoms, while number of incoming calls (calls/day: *β*=0.062, p=0.006), duration of incoming calls (seconds/day: *β*=30.38, p=0.011), number of outgoing calls (calls/day: *β*=0.15, p=0.006), and number of outgoing text messages (texts/day: *β*=0.24, p=0.034) was associated with manic symptoms [55]. In a separate cohort with additional digital biomarkers, Faurholt-Jepsen et al. (2016a) observed that during depressive states, individuals exhibited prolonged mobile phone screen-on times (seconds/day: *β*=209.6, p=0.04), received more incoming calls (calls/day: *β*=0.05, p=0.04), were less likely to answer incoming calls (calls/day: *β*=0.05, p=0.006), and made fewer outgoing calls (calls/day: *β*=-0.12, p=0.03) [25]. Conversely, during manic episodes, participants sent more outgoing text messages (texts/day: *β*=0.31, p=0.01), made longer phone calls (seconds/day: *β*=0.63, p=0.042), and received shorter text messages (characters/text: *β*=-3.54, p=0.001) [25].

Dominiak et al. (2022) built upon the results of the Faurholt-Jepsen et al. studies and found that depressed patients made less phone calls (calls/day: *β*=-0.064, p=0.01) and had a higher fraction of missed calls (*β*=4.43, p<0.001) compared to euthymia, while manic/mixed patients had more outgoing than incoming calls (*β*=2.73, p=0.03) compared to euthymia [56]. Additionally, Grünerbl et al. (2015) found that socialization metrics were an important marker in their multivariable Bayes classification algorithm and achieved an accuracy of 76% in predicting the current mood state using smartphone data and all sensor data [57]. Together, these studies indicate that increased social communication is linked to manic episodes, while reduced social interactions and communication avoidance are characteristic of depressive episodes.

#### Speech

Speech was studied in nine studies using voice recordings from real-world or clinical settings for mood state classification. Zhang et al. (2018) identified speech digital biomarkers, the fourth formant (p<0.05) and linear prediction coefficient (p<0.05), that decreased as patients transitioned from manic to euthymic states, and also found a significant correlation between the linear prediction coefficient and BRMS scores (r=0.398, p=0.040) [58]. Siegel et al. (2024) reported a positive correlation between speech latency and depression severity (r = 0.03, p<0.001) [20]. Another study explored demographic differences in speech-based digital biomarkers. Kaczmarek-Majer et al. (2024) demonstrated gender differences in speech digital biomarker associations with mania in terms of volume (male: *β*=1.6, female: *β*=-0.27), pitch (male: *β*=0.71, female: *β*=-0.21), and clarity (male: *β*=1.35, female: *β*=-0.25) [59]. Kaczmarek-Majer et al. (2024) also found male-specific speech correlations with bipolar depression in terms of volume (*β*=-1.07) and clarity (*β*=-1.00), but found no female-specific significant associations [59]. These studies demonstrate that various features of speech may reflect underlying mood states.

In addition to these associations, many studies developed speech classifiers to distinguish mood states with moderate to high accuracy. Ji et al. (2024) detected depressive and manic states from real-world “journal-style” voice recordings with high accuracy (AUROC=0.861 for depression and 0.903 for mania) [60]. Faurholt-Jepsen et al. (2016c) similarly found that voice digital biomarkers from phone calls were highly effective in classifying affective states, with an AUROC of 0.89 for mania and 0.78 for depression [61]. Karam et al. (2014) used speech recorded during clinical evaluations and built a classifier for manic (AUROC=0.81) and depressive states (AUROC=0.67) [19]. Gideon et al. (2016) also found that speech digital biomarkers derived from clinical interviews could reliably classify manic (AUROC=0.72±0.20) and depressive states (AUROC=0.75±0.14) [62]. Pan et al. (2018) differentiated speech by mood episodes in spontaneous speech in conversations with clinicians and found that support vector machines performed best for classification within individuals (AUROC=0.886), while Gaussian mixture models were superior for classification across individuals (AUROC=0.727) [63]. Together, speech features like pitch and speech latency offer a non-invasive and high-fidelity insight into bipolar mood dynamics, with growing evidence for their diagnostic and predictive validity.

### Multivariable Modeling

Seven studies utilized multiple categories of digital biomarkers extracted from wearable sensors and/or smartphones to classify mood states. Three of the seven studies came from the same group, and each took a slightly different approach toward developing deep learning models for the classification of mood states based on longitudinal wearable and smartphone data [64]. Across these studies, data were collected continuously for a minimum of 30 days in 37 to 149 BD patients and all analyses used digital biomarkers extracted from the Fitbit Charge wearable device and light exposure digital biomarkers extracted from Samsung smartphones. Cho et al. (2019) used 130 circadian-rhythm-based digital biomarkers in 37 BD patients as inputs to Random Forest models. The models were trained on data from the past 18 days to predict mood episodes in the following 3 days [34]. They reported an AUROC from 0.84 to 0.93 for classifying mood states (euthymia, depression, mania, and hypomania) and identified the most important digital biomarkers as steps during bedtime and daily light exposure for depression, sleep length and sleep quality for mania, and steps during bedtime and daily light exposure for hypomania [34]. Lee et al. (2023) extended the Random Forest mood episode prediction analysis to 175 BD patients and 140 features. They reported AUROC values from 0.95 to 0.98 for predicting depression, mania, and hypomania [35]. Step counts at bedtime (8-hour window before sunrise) was an important feature for predicting all three states [35]. Sleep efficiency was important for depression prediction, while morning light exposure and circadian rhythm acrophase/amplitude were important for (hypo)mania prediction [35]. The latest study published by this group used 36 digital biomarkers (all based on sleep and sleep-based circadian rhythms) and XGBoost to predict next-day mood episodes in 111 BD patients [54]. They found that circadian phase shifts (as estimated using the sleep/wake time series) were the most significant predictors across depressive, manic, and hypomanic episodes. Sleep/wake cycle delays were associated with depressive episodes, and advances were associated with manic episodes [54]. The XGBoost classifier yielded AUROC values of 0.925 to 0.985 for predicting next-day depression, mania, and hypomania [54].

The four remaining multivariable modeling studies use different approaches to classify mood states. Cote-Allard et al. (2022) proposed a novel deep learning-based ensemble method for euthymia/mania classification in 47 patients [23]. The authors used a deep learning model to classify mood states across patients with high accuracy (91.6%) based on 24-hour in-clinic recordings from the Empatica E4. The most important digital biomarkers for accurate classification were identified as acceleration and EDA [23]. Anmella et al. (2023) performed in-clinic recordings with the Empatica E4, but did so over a more extended time period of 48 hours and for repeated time points (acute episode, response, and remission) in eight patients [22]. Using a bidirectional long-short term memory neural network, they achieved 61-70% accuracy at classifying episode severity using acceleration, EDA, and HR digital biomarkers [22]. Evidence from Cote-Allard et al. (2022) and Anmella et al. (2023) converged on acceleration and EDA (via the Empatica E4) as the most important digital biomarkers in distinguishing mood episodes [23,65]. Grünerbl et al. (2015) was one of the earliest studies to apply multivariable ML techniques to classify mood states in real-world outpatients. The authors used digital biomarkers derived from smartphone socialization metrics, phone call speech characteristics, acceleration, and geolocation to classify states with 76% accuracy and a state change detection precision and recall of over 97% [57]. A more recent study by Lipschitz et al. (2024) used the Fitbit Inspire to monitor 17 digital biomarkers relevant to activity and sleep in 54 patients for 9 months. Their study used binary mixed model (BiMM) forest models to classify depression and (hypo)mania, with AUROC values of 0.86 for depression and 0.852 for (hypo)mania [36]. The most important digital biomarkers for depression classification were time awake at night, total sleep duration, median bedtime, and resting heart rate [36]. The most important digital biomarkers for (hypo)mania classification were heart rate, sleep efficiency, REM sleep percentage, and active minutes [36].

### Risk of Bias Analysis

#### PROBAST

14 studies were evaluated for the risk of bias in their development of predictive models using PROBAST [17]. While all studies were evaluated to have low risk of bias regarding participants, predictors, and outcomes, all studies demonstrated at least some risk in the analysis domain. This risk was primarily due to the low sample and event sizes and the highly complex multivariate models driving high risk regarding the model’s generalizability to broader populations. Additionally, many models were highly complex and were largely unexplainable in terms of which digital biomarkers were driving classification performance. In addition, none of the studies validated their models on an external dataset, proving that all studies included are somewhat biased and should be seen as preliminary. The complete PROBAST analysis is in Supplementary Table 2.

#### Newcastle-Ottawa

29 studies were evaluated for risk of bias using the Newcastle-Ottawa Scale (NOS), a widely used tool for assessing the quality of non-randomized studies [18]. Most studies (24 out of 29) were rated as high-quality with a low risk of bias as they showed strong methodological rigor in the selection of study groups, comparability, and ascertainment of either exposure or outcome. They typically included well-defined cohorts, appropriate follow-up periods, and robust outcome assessment procedures. However, a smaller subset of the studies (5 out of 29) was classified as moderate in quality and had some risk of bias. These studies often had missing points in the comparability domain, suggesting a lack of control for confounding factors (i.e., gender, race), or fewer strengths in outcome ascertainment or follow-up adequacy. Additionally, none of the 29 studies received points for comparability of cohorts against a control group, as it was not the focus of many of the studies. The complete Newcastle-Ottawa assessment is presented in Supplementary Table 3.

## Discussion

### Principal Findings

This systematic review identified 45 studies meeting our inclusion criteria that collected objective data from remote monitoring devices (e.g. smartphone and wearable sensors) and found strong associations with BD mood states. Specifically, we found nine categories of objective digital biomarkers: physical activity, HR, EDA, geolocation, keyboard usage, light exposure, sleep, socialization, and speech. While several digital biomarkers were consistently associated with mood states (e.g., decreased geographic movements and smartphone socialization in depression), evidence for others was mixed (e.g., heart rate variability in mania). While these studies demonstrate early promise, their heterogeneity in design, duration, and analysis highlights the need for stronger methodological consensus and clinical validation. The converging evidence across studies points toward a future where objective, real-time digital biomarkers may not only detect mood transitions but also guide targeted interventions.

### Diagnostically Grounded vs. Exploratory Digital Biomarkers

Five of the digital marker categories we identified are directly grounded in the diagnostic criteria of BD: geolocation, physical activity, sleep, socialization, and speech. These behaviors correspond to the energy and social engagement dimensions of manic and depressive episodes, as specified in the DSM-5 [66]. The DSM-5 outlines increased goal-directed activity, decreased need for sleep, and heightened sociability as core characteristics of manic episodes, while hypersomnia and social withdrawal are typical of depressive episodes. The alignment of these behaviors with digital biomarkers strengthens their clinical validity. For example, manic states were associated with increased activity [25,50], shorter and more frequent phone calls [25], and shifts in sleep patterns [54], while depressive states were accompanied by decreased movement [48], reduced call frequency [56], and increased sleep variability [52]. Additionally, the DSM-5 recognizes circadian rhythm disruption as a clinically significant marker in BD. Some digital biomarkers in the physical activity, light exposure, and sleep categories can also be classified as circadian rhythm-based digital biomarkers. Specifically, circadian phase, amplitude based on the sleep-wake time series [54], and steps during nighttime [34,35] were identified as important digital biomarkers in deep-learning algorithms for mood state classification. Additionally, circadian activity rhythms based on accelerometer measurements have significant associations with mood episode relapses: specifically, robust circadian activity rhythms were associated with a decrease in mood episode relapses, and a later timing was associated with depressive episode relapses [49]. These findings are directly aligned with the understanding that circadian rhythm dysfunction has long been associated with BD [67].

In contrast, digital biomarkers like EDA, heart rate variability, keyboard usage, and light exposure are more exploratory and less tied to DSM criteria, but may reflect latent physiological processes underlying mood dysregulation. Specifically, EDA may provide insight into autonomic nervous system functioning and has been found to be consistently lower in those with depression [68]. In addition, bedroom light exposure may be an indirect measure of mania [41] or future manic episodes [42], while daytime light exposure intensity may indicate fewer depressive symptoms [43] and episode relapses [44]. Longitudinal studies in larger cohorts will reveal the consistency and generalizability of these exploratory digital biomarkers as predictors of mood state.

### Need for Longitudinal Monitoring

A critical methodological gap identified in this review and reinforced through our bias analyses is the mismatch between the duration of monitoring and episodic nature of BD. Mood episodes often unfold over weeks to months, yet many studies implemented monitoring windows too brief to capture meaningful mood transitions. For example, studies by Pan et al. (2018), Cote-Allard et al. (2022), and Wazen et al. (2018) collected data for less than a month in primarily inpatient environments, limiting the ability to detect the naturalistic dynamics of mood changes [23,30,63]. Additionally, for studies that aim to identify markers of hypomanic and mixed states, it is challenging to predict mood states daily when activity is collected per minute and symptoms are evaluated monthly [48]. In contrast, the most informative and clinically promising studies were those with deployed longitudinal protocols. Lipschitz et al. (2024) followed participants over nine months while Lim et al. (2024) analyzed 260 days of continuous wearable data [36,54]. Both studies captured multiple mood episodes and transitions, allowing for the development of temporally rich prediction models. Similarly, the APPLE cohort studies used long-term monitoring to understand circadian rhythm fluctuations [41]. These extended durations allow for robust within-subject modeling, which is critical for identifying pre-episode changes. Future work must prioritize long-term, continuous data collection to enable predictive modeling of transitions into and out of clinically relevant states.

### Frequency of Clinical Labels

To build clinically useful models, the frequency of ground truth labels must align with both the temporal resolution of passive data monitoring and the clinical validity of assessment tools. Although scales commonly administered in these studies, like the YMRS or MADRS, are typically designed to be administered every 2 weeks, many studies chose longer intervals (up to three months or longer), compromising the ability to capture transient states. For example, several studies used monthly or even less frequent mood assessments that could miss shorter-lived episodes or rapid cycling [62,63]. By contrast, some studies administered bi-weekly clinical scales, striking a better balance between patient burden and temporal precision [24,36]. In the absence of frequent clinical touchpoints, many studies relied on retrospective labeling of mood states using the most recent clinician-administered scale. The retroactive labeling potentially introduces uncertainty when trying to link mood states to passive data on a day-to-day basis, especially when mood fluctuates rapidly or data collection is continuous. Studies should be designed to administer validated clinical scales at recommended intervals to maximize temporal alignment with passive data and enhance the reliability of supervised learning models. Future studies should explicitly define their protocols for labeling ground truth mood states and report how clinician-rated scores are temporally mapped to passive sensing windows, as these decisions critically affect downstream model performance.

### Machine Learning Methods for Mood State Classification

The recent emergence of ML and AI, present in 15 included studies, marks a significant shift toward more sophisticated monitoring practices. This emergence is well-documented, with other recent reviews covering the application of ML and AI-based approaches in detecting mental health states [69,70]. These recent reviews come to a similar conclusion as this review: further work is needed to validate findings [69,70]. While many studies reported high classification accuracy, few explored the trade-offs between sensitivity and specificity, which are critical for patient care. False positives may lead to over-monitoring and increased patient burden, whereas false negatives may delay care. Model optimization should also consider the deployment context and temporal nature of the data collected when recommending an intervention. Common pitfalls included overfitting due to small sample sizes, lack of interpretability, and temporally unaware models. No included studies combined more than four marker categories we identified in predictive models. In the future, researchers should consider a smartphone and wearable-based approach to passive sensing that integrates the digital biomarkers summarized in this review for improved predictive performance, as wearable-only sensing approaches miss out on important smartphone-based digital biomarkers that have shown to be implicated in BD such as geolocation and socialization.

### Need for Objective Biomarkers to Understand Disease Mechanisms

Psychiatric care has long relied on observed behaviors and self-reported symptoms, which are intermittent, context-dependent, and only indirectly tied to underlying biology. Reliance on behaviors and symptoms limits mechanistic insight and makes it difficult to build objective markers that generalize across patients and settings. In our review, only two digital biomarker categories (HR and EDA) were primarily physiological, while the rest capture behavioral manifestations of illness. This lack of biological and physiological markers is mainly due to the limited capability of widely accessible wearable sensors to capture relevant signals. To move the field toward a biologically grounded understanding of bipolar disorder, future work should pair continuous behavioral sensing with equally continuous measures of physiology and neural activity. Practical avenues include emerging biochemical and physiological sensors, including skin patches that sample biochemical signals such as stress hormones or inflammatory markers [71], and recording-capable neuromodulation devices like deep brain stimulation systems used in exploratory psychiatric research [72,73]. Prospective studies should collect these streams in the same individuals over extended periods, align them to clearly defined clinical events, and test whether biological signals can explain clinical states beyond behavior alone. In the end, a map linking behavior, physiology, biochemistry, and brain activity would clarify mechanisms of psychiatric disorders and identify actionable targets.

### Ethical Considerations

The limited validity, accuracy, generalizability, and reliability of existing digital biomarkers for BD and other conditions carry well-recognized ethical ramifications because they directly affect the efficacy and safety of biomarker-guided clinical decisions. Yet, concerns extend beyond the predictive accuracy of mood states. Efforts to identify objective digital biomarkers for BD also raise serious privacy issues: passively and continuously capturing multiple streams of sensitive data as patients navigate their daily lives can expose intimate details of behavior and physiology [74]. Consequently, we advocate a data minimization approach that restricts collection to streams with clear, empirically demonstrated clinical value—that is, data shown to meaningfully refine assessments of a patient’s health status or to inform treatment choices[75,76]. Future research should aim to converge on a smaller, purpose-driven subset of digital biomarkers that demonstrably predict clinically relevant states. This principled narrowing is essential both to protect patient privacy and to ensure that only the most informative digital biomarkers enter routine clinical practice.

### Missing Data and Compliance

Missing data is a pervasive issue and is closely tied to patient compliance and clinical status. In the included studies, there is also a lack of reporting and variability on the handling of missing data and data quality, which is a major concern when it comes to smartphone and wearable data. Few studies explicitly modeled missingness, but several [36,77] acknowledged gaps in wearable device usage. In our own work in patients with obsessive-compulsive disorder (OCD), we found that patients were more likely to engage in remote monitoring activities (e.g., recharging wearable devices, responding to self-reports) when OCD symptoms were minimal, suggesting that lack of compliance may indicate symptom exacerbations [72]. Likewise, non-wear during depressive episodes could signal social withdrawal or fatigue, whereas erratic device usage might suggest mania. Rather than ignoring missing data patterns or treating them as noise, future models may consider integrating missing data as a potentially informative signal for this population [78]. To improve data completeness and clinical utility, it may be best to design passive sensing systems that reduce the need for active participation. Passive sensing would reduce patient burden and bypass states characterized by poor insight when patients cannot identify warning signs in their behavior. Reducing obtrusiveness and invasiveness not only enhances scalability but also supports ethical goals like patient autonomy, promoting ease of use, and elevates sensing to the status of a potential pre-episode clinical marker..

### Operationalizing Digital Biomarkers for Clinical Use

Moving beyond passive monitoring towards meaningful clinical use requires digital biomarkers to become actionable tools that can inform treatment decisions. Passive signals must first be validated as reliable indicators of mood state changes across diverse and representative populations. Several studies in this review demonstrated the potential for mood state classification [36,54], but few linked their results to concrete clinical actions. To be actionable, there must be predefined interventions for each digital biomarker, such as clinician alerts or medication adjustments. There will need to be extensive collaboration between data scientists, clinicians, ethicists, and possibly participatory approaches with patients to establish clinically meaningful thresholds for action (e.g. how much of a deviation in heart rate variability warrants a clinical intervention). Real-time monitoring must be supported by an infrastructure that enables clinicians to integrate these interventions into care delivery systems in a minimally invasive way, balancing accuracy and avoiding overburdening of patients or clinicians [70]. Overall, utilizing digital biomarkers for intervention will require technical precision and careful integration into existing healthcare systems.

### Limitations of this Review

We chose to exclude studies that focused on unipolar depression and/or entangled BD results with other disorders. While the exclusion allowed for a more focused review, it may have omitted relevant findings or generalizable methods that could inform BD-specific monitoring strategies. Other related reviews solely focus on monitoring depression [69], suicidality [79], or psychosis [80]. We chose to include some studies with methods that are not directly possible for remote monitoring (e.g., external photometers and microphones) but could be made possible using alternative sensors (e.g., via smartphone or wearable microphones). Additionally, we identified bias in nearly all studies due to low sample and event sizes, especially studies that developed predictive models (see Supplementary Table 3). Hypomanic/manic episodes meeting diagnostic criteria may be less frequent than sub-threshold episodes and are therefore difficult to capture outside of the inpatient setting [1]. However, the inpatient setting lacks ethological validity. Therefore, the digital biomarkers presented in this review need to be further validated in larger and more diverse cohorts with longer and more frequent clinical touchpoints. Heterogeneity in patient follow-up, measurement protocols, and reporting of results may limit the reproducibility and comparison of findings across studies. For example, six different types of wearables used in the included studies, as well as different types of smartphones (Android, Apple). Different devices often use proprietary algorithms to derive behavioral and physiological features, like sleep staging, leading to inconsistent outputs [81].

### Conclusion and Future Directions

Digital biomarkers derived from smartphone and wearable sensors present a promising pathway toward precision psychiatry in BD, enabling continuous, real-time monitoring of mood states. This review highlights several categories of digital biomarkers that appear clinically promising, such as physical activity, sleep, and speech. In particular, markers tied to circadian rhythms (nighttime light usage and sleep-wake cycles) have demonstrated robust predictive power across studies and align with diagnostic criteria. Researchers and clinicians need to further establish which objective digital biomarkers have clinical significance to improve psychiatric practice through longitudinal clinical trials in larger cohorts. Next steps should include standardizing features across devices, validating models across diverse populations, and integrating passive monitoring into clinical workflows. Ethical implementation will also require balancing privacy with clinical actionability. ML and AI models for predicting mood states are promising but need to be further validated, and can be leveraged in the future to implement the digital biomarkers summarized in this review for improved performance. Remote monitoring has the potential to fundamentally shift the clinical management of BD by enabling earlier detection of mood state transitions and real-time risk assessment, revolutionizing how psychiatric care is delivered.

## Author Contributions

N.R.P., S.A.S., J.A.H., W.K.G., T.P.K., and I.C. devised the study. T.P.K. and I.C. devised the search terms and inclusion/exclusion criteria, conducted the searches, screened the abstracts and full papers, extracted the data and performed the risk-of-bias/quality appraisals, and carried out the synthesis. T.P.K. and I.C. wrote the paper, which was reviewed and approved by all authors (K.K.Q., A.San., N.M., J.A.H., W.K.G., S.A.S., A.Sab., N.R.P.).

## Acknowledgements

This research was supported by the National Institutes of Health (NIH) National Institute of Neurological Disorders and Stroke BRAIN Initiative via contract UH3NS136631 (to W.K.G., S.A.S., J.A.H., N.R.P.). This material is based upon work supported by the National Science Foundation Graduate Research Fellowship Program under Grant No. 1842494 (T.P.K.). The authors would also like to acknowledge support from a fellowship from the Gulf Coast Consortia on the NLM Training Program in Biomedical Informatics and Data Science T15LM007093 (I.C.).

## Data Availability

All data used in this study are reported in the main text and Supplementary Materials.

## References

1. Grande I, Berk M, Birmaher B, Vieta E. Bipolar disorder. The Lancet Elsevier; 2016 Apr 9;387(10027):1561–1572. PMID:26388529

2. Vieta E, Berk M, Schulze TG, Carvalho AF, Suppes T, Calabrese JR, Gao K, Miskowiak KW, Grande I. Bipolar disorders. Nat Rev Dis Primer 2018 Mar 8;4(1):18008. doi: 10.1038/nrdp.2018.8

3. Judd LL, Akiskal HS, Schettler PJ, Endicott J, Maser J, Solomon DA, Leon AC, Rice JA, Keller MB. The long-term natural history of the weekly symptomatic status of bipolar I disorder. Arch Gen Psychiatry 2002 June;59(6):530–537. PMID:12044195

4. Judd LL, Akiskal HS, Schettler PJ, Coryell W, Endicott J, Maser JD, Solomon DA, Leon AC, Keller MB. A prospective investigation of the natural history of the long-term weekly symptomatic status of bipolar II disorder. Arch Gen Psychiatry 2003 Mar;60(3):261–269. PMID:12622659

5. Simon GE, Hunkeler E, Fireman B, Lee JY, Savarino J. Risk of suicide attempt and suicide death in patients treated for bipolar disorder. Bipolar Disord 2007;9(5):526–530. doi: 10.1111/j.1399-5618.2007.00408.x

6. Plans L, Barrot C, Nieto E, Rios J, Schulze TG, Papiol S, Mitjans M, Vieta E, Benabarre A. Association between completed suicide and bipolar disorder: A systematic review of the literature. J Affect Disord 2019 Jan 1;242:111–122. doi: 10.1016/j.jad.2018.08.054

7. Ortiz A, Mulsant BH. Beyond Step Count: Are We Ready to Use Digital Phenotyping to Make Actionable Individual Predictions in Psychiatry? J Med Internet Res 2024 Aug 5;26:e59826. doi: 10.2196/59826

8. Bauer M, Andreassen OA, Geddes JR, Vedel Kessing L, Lewitzka U, Schulze TG, Vieta E. Areas of uncertainties and unmet needs in bipolar disorders: clinical and research perspectives. Lancet Psychiatry 2018 Nov 1;5(11):930–939. doi: 10.1016/S2215-0366(18)30253-0

9. Faurholt-Jepsen M, Frost M, Ritz C, Christensen EM, Jacoby AS, Mikkelsen RL, Knorr U, Bardram JE, Vinberg M, Kessing LV. Daily electronic self-monitoring in bipolar disorder using smartphones – the MONARCA I trial: a randomized, placebo-controlled, single-blind, parallel group trial. Psychol Med 2015 Oct;45(13):2691–2704. doi: 10.1017/S0033291715000410

10. Faurholt-Jepsen M, Frost M, Christensen EM, Bardram JE, Vinberg M, Kessing LV. The effect of smartphone-based monitoring on illness activity in bipolar disorder: the MONARCA II randomized controlled single-blinded trial. Psychol Med 2020 Apr;50(5):838–848. doi: 10.1017/S0033291719000710

11. Peralta V, Cuesta MJ. Lack of insight in mood disorders. J Affect Disord 1998 Apr;49(1):55–58. PMID:9574860

12. Saccaro LF, Amatori G, Cappelli A, Mazziotti R, Dell’Osso L, Rutigliano G. Portable technologies for digital phenotyping of bipolar disorder: A systematic review. J Affect Disord 2021 Dec 1;295:323–338. doi: 10.1016/j.jad.2021.08.052

13. Ortiz A, Maslej MM, Husain MI, Daskalakis ZJ, Mulsant BH. Apps and gaps in bipolar disorder: A systematic review on electronic monitoring for episode prediction. J Affect Disord 2021 Dec 1;295:1190–1200. PMID:34706433

14. Moher D, Liberati A, Tetzlaff J, Altman DG, The PRISMA Group. Preferred Reporting Items for Systematic Reviews and Meta-Analyses: The PRISMA Statement. PLoS Med 2009 July 21;6(7):e1000097. doi: 10.1371/journal.pmed.1000097

15. Vasudevan S, Saha A, Tarver ME, Patel B. Digital biomarkers: Convergence of digital health technologies and biomarkers. Npj Digit Med Nature Publishing Group; 2022 Mar 25;5(1):1–3. doi: 10.1038/s41746-022-00583-z

16. Ouzzani M, Hammady H, Fedorowicz Z, Elmagarmid A. Rayyan—a web and mobile app for systematic reviews. Syst Rev 2016 Dec 5;5(1):210. doi: 10.1186/s13643-016-0384-4

17. Moons KGM, Wolff RF, Riley RD, Whiting PF, Westwood M, Collins GS, Reitsma JB, Kleijnen J, Mallett S. PROBAST: A Tool to Assess Risk of Bias and Applicability of Prediction Model Studies: Explanation and Elaboration. Ann Intern Med American College of Physicians; 2019 Jan;170(1):W1–W33. doi: 10.7326/M18-1377

18. Wells G, Shea B, O’Connel D, Peterson J, Welch V, Losos M, Tugwell P. The Newcastle-Ottawa Scale (NOS) for assessing the quality of nonrandomised studies in meta-analyses. 2000. Available from: https://www.ohri.ca/programs/clinical_epidemiology/oxford.asp [accessed Feb 28, 2025]

19. Karam ZN, Provost EM, Singh S, Montgomery J, Archer C, Harrington G, Mcinnis MG. Ecologically valid long-term mood monitoring of individuals with bipolar disorder using speech. 2014 IEEE Int Conf Acoust Speech Signal Process ICASSP Florence, Italy: IEEE; 2014. p. 4858–4862. doi: 10.1109/ICASSP.2014.6854525

20. Siegel JS, Cohen AS, Szabo ST, Tomioka S, Opler M, Kirkpatrick B, Hopkins S. Enrichment using speech latencies improves treatment effect size in a clinical trial of bipolar depression. Psychiatry Res 2024 Oct;340:116105. PMID:39151277

21. Valenza G, Nardelli M, Lanata A, Gentili C, Bertschy G, Paradiso R, Scilingo EP. Wearable Monitoring for Mood Recognition in Bipolar Disorder Based on History-Dependent Long-Term Heart Rate Variability Analysis. IEEE J Biomed Health Inform 2014 Sept;18(5):1625–1635. doi: 10.1109/JBHI.2013.2290382

22. Anmella G, Corponi F, Li BM, Mas A, Sanabra M, Pacchiarotti I, Valentí M, Grande I, Benabarre A, Giménez-Palomo A, Garriga M, Agasi I, Bastidas A, Cavero M, Fernández-Plaza T, Arbelo N, Bioque M, García-Rizo C, Verdolini N, Madero S, Murru A, Amoretti S, Martínez-Aran A, Ruiz V, Fico G, De Prisco M, Oliva V, Solanes A, Radua J, Samalin L, Young AH, Vieta E, Vergari A, Hidalgo-Mazzei D. Exploring Digital Biomarkers of Illness Activity in Mood Episodes: Hypotheses Generating and Model Development Study. JMIR MHealth UHealth 2023 May 4;11:e45405. doi: 10.2196/45405

23. Côté Allard U, Jakobsen P, Stautland A, Nordgreen T, Fasmer OB, Oedegaard K, Torresen J. Long–Short Ensemble Network for Bipolar Manic-Euthymic State Recognition Based on Wrist-Worn Sensors. IEEE Pervasive Comput 2022 Apr 4;1–12. doi: 10.1109/MPRV.2022.3155728

24. Faurholt-Jepsen M, Frost M, Vinberg M, Christensen EM, Bardram JE, Kessing LV. Smartphone data as objective measures of bipolar disorder symptoms. Psychiatry Res 2014 June 30;217(1–2):124–127. PMID:24679993

25. Faurholt-Jepsen M, Vinberg M, Frost M, Debel S, Margrethe Christensen E, Bardram JE, Kessing LV. Behavioral activities collected through smartphones and the association with illness activity in bipolar disorder. Int J Methods Psychiatr Res 2016 Apr 1;25(4):309–323. PMID:27038019

26. Faurholt-Jepsen M, Busk J, Vinberg M, Christensen EM, HelgaÞórarinsdóttir null, Frost M, Bardram JE, Kessing LV. Daily mobility patterns in patients with bipolar disorder and healthy individuals. J Affect Disord 2021 Jan 1;278:413–422. PMID:33010566

27. Palmius N, Tsanas A, Saunders KEA, Bilderbeck AC, Geddes JR, Goodwin GM, De Vos M. Detecting Bipolar Depression From Geographic Location Data. IEEE Trans Biomed Eng 2017 Aug;64(8):1761–1771. PMID:28113247

28. Beiwinkel T, Kindermann S, Maier A, Kerl C, Moock J, Barbian G, Rössler W. Using Smartphones to Monitor Bipolar Disorder Symptoms: A Pilot Study. JMIR Ment Health 2016 Jan 6;3(1):e2. doi: 10.2196/mental.4560

29. Faurholt-Jepsen M, Brage S, Kessing LV, Munkholm K. State-related differences in heart rate variability in bipolar disorder. J Psychiatr Res 2017 Jan;84:169–173. PMID:27743529

30. Wazen GLL, Gregório ML, Kemp AH, Godoy MFD. Heart rate variability in patients with bipolar disorder: From mania to euthymia. J Psychiatr Res 2018 Apr;99:33–38. doi: 10.1016/j.jpsychires.2018.01.008

31. Corponi F, Li BM, Anmella G, Valenzuela-Pascual C, Pacchiarotti I, Valentí M, Grande I, Benabarre A, Garriga M, Vieta E, Lawrie SM, Whalley HC, Hidalgo-Mazzei D, Vergari A. A Bayesian analysis of heart rate variability changes over acute episodes of bipolar disorder. Npj Ment Health Res 2024 Oct 3;3(1):44. doi: 10.1038/s44184-024-00090-x

32. Valenza G, Nardelli M, Lanata A, Gentili C, Bertschy G, Kosel M, Scilingo EP. Predicting Mood Changes in Bipolar Disorder Through Heartbeat Nonlinear Dynamics. IEEE J Biomed Health Inform 2016 July;20(4):1034–1043. doi: 10.1109/JBHI.2016.2554546

33. Nardelli M, Lanata A, Bertschy G, Scilingo EP, Valenza G. Heartbeat Complexity Modulation in Bipolar Disorder during Daytime and Nighttime. Sci Rep 2017 Dec 20;7(1):17920. doi: 10.1038/s41598-017-18036-z

34. Cho C-H, Lee T, Kim M-G, In HP, Kim L, Lee H-J. Mood Prediction of Patients With Mood Disorders by Machine Learning Using Passive Digital Phenotypes Based on the Circadian Rhythm: Prospective Observational Cohort Study. J Med Internet Res 2019 Apr 17;21(4):e11029. doi: 10.2196/11029

35. Lee H-J, Cho C-H, Lee T, Jeong J, Yeom JW, Kim S, Jeon S, Seo JY, Moon E, Baek JH, Park DY, Kim SJ, Ha TH, Cha B, Kang H-J, Ahn Y-M, Lee Y, Lee J-B, Kim L. Prediction of impending mood episode recurrence using real-time digital phenotypes in major depression and bipolar disorders in South Korea: a prospective nationwide cohort study. Psychol Med 2023 Sept;53(12):5636–5644. doi: 10.1017/S0033291722002847

36. Lipschitz JM, Lin S, Saghafian S, Pike CK, Burdick KE. Digital phenotyping in bipolar disorder: Using longitudinal Fitbit data and personalized machine learning to predict mood symptomatology. Acta Psychiatr Scand 2024 Oct 13;acps.13765. doi: 10.1111/acps.13765

37. Liu Q, Ning E, Ross MK, Cladek A, Kabir S, Barve A, Kennelly E, Hussain F, Duffecy J, Langenecker SA, Nguyen TM, Tulabandhula T, Zulueta J, Demos AP, Leow A, Ajilore O. Digital Phenotypes of Mobile Keyboard Backspace Rates and Their Associations With Symptoms of Mood Disorder: Algorithm Development and Validation. J Med Internet Res 2024 Oct 29;26:e51269. doi: 10.2196/51269

38. Stange JP, Zulueta J, Langenecker SA, Ryan KA, Piscitello A, Duffecy J, McInnis MG, Nelson P, Ajilore O, Leow A. Let your fingers do the talking: Passive typing instability predicts future mood outcomes. Bipolar Disord 2018 May;20(3):285–288. doi: 10.1111/bdi.12637

39. Zulueta J, Piscitello A, Rasic M, Easter R, Babu P, Langenecker SA, McInnis M, Ajilore O, Nelson PC, Ryan K, Leow A. Predicting Mood Disturbance Severity with Mobile Phone Keystroke Metadata: A BiAffect Digital Phenotyping Study. J Med Internet Res 2018 July 20;20(7):e241. doi: 10.2196/jmir.9775

40. Cao B, Zheng L, Zhang C, Yu PS, Piscitello A, Zulueta J, Ajilore O, Ryan K, Leow AD. DeepMood: Modeling Mobile Phone Typing Dynamics for Mood Detection. Proc 23rd ACM SIGKDD Int Conf Knowl Discov Data Min Halifax NS Canada: ACM; 2017. p. 747–755. doi: 10.1145/3097983.3098086

41. Esaki Y, Obayashi K, Saeki K, Fujita K, Iwata N, Kitajima T. Association between light exposure at night and manic symptoms in bipolar disorder: cross-sectional analysis of the APPLE cohort. Chronobiol Int 2020 June;37(6):887–896. PMID:32238002

42. Esaki Y, Obayashi K, Saeki K, Fujita K, Iwata N, Kitajima T. Effect of nighttime bedroom light exposure on mood episode relapses in bipolar disorder. Acta Psychiatr Scand 2022 July;146(1):64–73. PMID:35253206

43. Esaki Y, Kitajima T, Obayashi K, Saeki K, Fujita K, Iwata N. Daytime light exposure in daily life and depressive symptoms in bipolar disorder: A cross-sectional analysis in the APPLE cohort. J Psychiatr Res 2019 Sept;116:151–156. PMID:31247358

44. Esaki Y, Obayashi K, Saeki K, Fujita K, Iwata N, Kitajima T. Preventive effect of morning light exposure on relapse into depressive episode in bipolar disorder. Acta Psychiatr Scand 2021 Apr;143(4):328–338. PMID:33587769

45. Cho C-H, Ahn Y-M, Kim SJ, Ha TH, Jeon HJ, Cha B, Moon E, Park DY, Baek JH, Kang H-J, Ryu V, An H, Lee H-J. Design and Methods of the Mood Disorder Cohort Research Consortium (MDCRC) Study. Psychiatry Investig 2017 Jan;14(1):100–106. PMID:28096882

46. Ebner-Priemer UW, Mühlbauer E, Neubauer AB, Hill H, Beier F, Santangelo PS, Ritter P, Kleindienst N, Bauer M, Schmiedek F, Severus E. Digital phenotyping: towards replicable findings with comprehensive assessments and integrative models in bipolar disorders. Int J Bipolar Disord 2020 Dec;8(1):35. doi: 10.1186/s40345-020-00210-4

47. Faurholt-Jepsen M, Brage S, Vinberg M, Kessing LV. State-related differences in the level of psychomotor activity in patients with bipolar disorder - Continuous heart rate and movement monitoring. Psychiatry Res 2016 Mar 30;237:166–174. PMID:26832835

48. Gershon A, Ram N, Johnson SL, Harvey AG, Zeitzer JM. Daily Actigraphy Profiles Distinguish Depressive and Interepisode States in Bipolar Disorder. Clin Psychol Sci J Assoc Psychol Sci 2016 July;4(4):641–650. PMID:27642544

49. Esaki Y, Obayashi K, Saeki K, Fujita K, Iwata N, Kitajima T. Association between circadian activity rhythms and mood episode relapse in bipolar disorder: a 12-month prospective cohort study. Transl Psychiatry Nature Publishing Group; 2021 Oct 13;11(1):1–8. doi: 10.1038/s41398-021-01652-9

50. Jakobsen P, Stautland A, Riegler MA, Côté-Allard U, Sepasdar Z, Nordgreen T, Torresen J, Fasmer OB, Oedegaard KJ. Complexity and variability analyses of motor activity distinguish mood states in bipolar disorder. PLOS ONE Public Library of Science; 2022 Jan 21;17(1):e0262232. doi: 10.1371/journal.pone.0262232

51. Ortiz A, Halabi R, Alda M, Burgos A, DeShaw A, Gonzalez-Torres C, Husain MI, O’Donovan C, Tolend M, Hintze A, Mulsant BH. Day-to-day variability in sleep and activity predict the onset of a hypomanic episode in patients with bipolar disorder. J Affect Disord 2025 Apr;374:75–83. doi: 10.1016/j.jad.2025.01.026

52. Esaki Y, Obayashi K, Saeki K, Fujita K, Iwata N, Kitajima T. Circadian variability of objective sleep measures predicts the relapse of a mood episode in bipolar disorder: findings from the APPLE cohort. Psychiatry Clin Neurosci 2023 Aug;77(8):442–448. doi: 10.1111/pcn.13556

53. Esaki Y, Obayashi K, Saeki K, Fujita K, Iwata N, Kitajima T. Daytime napping and depressive symptoms in bipolar disorder: A cross-sectional analysis of the APPLE cohort. Sleep Med 2024 Dec 1;124:688–694. doi: 10.1016/j.sleep.2024.11.006

54. Lim D, Jeong J, Song YM, Cho C-H, Yeom JW, Lee T, Lee J-B, Lee H-J, Kim JK. Accurately predicting mood episodes in mood disorder patients using wearable sleep and circadian rhythm features. Npj Digit Med 2024 Nov 18;7(1):324. doi: 10.1038/s41746-024-01333-z

55. Faurholt-Jepsen M, Vinberg M, Frost M, Christensen EM, Bardram JE, Kessing LV. Smartphone data as an electronic biomarker of illness activity in bipolar disorder. Bipolar Disord 2015 Nov;17(7):715–728. PMID:26395972

56. Dominiak M, Kaczmarek-Majer K, Antosik-Wójcińska AZ, Opara KR, Olwert A, Radziszewska W, Hryniewicz O, Święcicki Ł, Wojnar M, Mierzejewski P. Behavioral and Self-reported Data Collected From Smartphones for the Assessment of Depressive and Manic Symptoms in Patients With Bipolar Disorder: Prospective Observational Study. J Med Internet Res 2022 Jan 19;24(1):e28647. PMID:34874015

57. Grunerbl A, Muaremi A, Osmani V, Bahle G, Ohler S, Troster G, Mayora O, Haring C, Lukowicz P. Smartphone-Based Recognition of States and State Changes in Bipolar Disorder Patients. IEEE J Biomed Health Inform 2015 Jan;19(1):140–148. doi: 10.1109/JBHI.2014.2343154

58. Zhang J, Pan Z, Gui C, Xue T, Lin Y, Zhu J, Cui D. Analysis on speech signal features of manic patients. J Psychiatr Res 2018 Mar;98:59–63. doi: 10.1016/j.jpsychires.2017.12.012

59. Kaczmarek-Majer K, Dominiak M, Antosik AZ, Hryniewicz O, Kamińska O, Opara K, Owsiński J, Radziszewska W, Sochacka M, Święcicki Ł. Acoustic features from speech as markers of depressive and manic symptoms in bipolar disorder: A prospective study. Acta Psychiatr Scand 2024 Aug 8;acps.13735. doi: 10.1111/acps.13735

60. Ji J, Dong W, Li J, Peng J, Feng C, Liu R, Shi C, Ma Y. Depressive and mania mood state detection through voice as a biomarker using machine learning. Front Neurol 2024 July 4;15:1394210. doi: 10.3389/fneur.2024.1394210

61. Faurholt-Jepsen M, Busk J, Frost M, Vinberg M, Christensen EM, Winther O, Bardram JE, Kessing LV. Voice analysis as an objective state marker in bipolar disorder. Transl Psychiatry 2016 July 19;6(7):e856–e856. doi: 10.1038/tp.2016.123

62. Gideon J, Provost EM, McInnis M. Mood state prediction from speech of varying acoustic quality for individuals with bipolar disorder. 2016 IEEE Int Conf Acoust Speech Signal Process ICASSP 2016. p. 2359–2363. doi: 10.1109/ICASSP.2016.7472099

63. Pan Z, Gui C, Zhang J, Zhu J, Cui D. Detecting Manic State of Bipolar Disorder Based on Support Vector Machine and Gaussian Mixture Model Using Spontaneous Speech. Psychiatry Investig 2018 July;15(7):695–700. PMID:29969852

64. Cho C-H, Ahn Y-M, Kim SJ, Ha TH, Jeon HJ, Cha B, Moon E, Park DY, Baek JH, Kang H-J, Ryu V, An H, Lee H-J. Design and Methods of the Mood Disorder Cohort Research Consortium (MDCRC) Study. Psychiatry Investig 2017 Jan;14(1):100–106. PMID:28096882

65. Anmella G, Corponi F, Li BM, Mas A, Sanabra M, Pacchiarotti I, Valentí M, Grande I, Benabarre A, Giménez-Palomo A, Garriga M, Agasi I, Bastidas A, Cavero M, Fernández-Plaza T, Arbelo N, Bioque M, García-Rizo C, Verdolini N, Madero S, Murru A, Amoretti S, Martínez-Aran A, Ruiz V, Fico G, Prisco MD, Oliva V, Solanes A, Radua J, Samalin L, Young AH, Vieta E, Vergari A, Hidalgo-Mazzei D. Exploring Digital Biomarkers of Illness Activity in Mood Episodes: Hypotheses Generating and Model Development Study. JMIR MHealth UHealth 2023 May 4;11(1):e45405. doi: 10.2196/45405

66. American Psychiatric Association. Diagnostic and Statistical Manual of Mental Disorders. 5th ed. Arlington, VA: American Psychiatric Publishing; 2013.

67. Takaesu Y. Circadian rhythm in bipolar disorder: A review of the literature. Psychiatry Clin Neurosci 2018;72(9):673–682. doi: 10.1111/pcn.12688

68. Thorell LH, Kjellman BF, D’Elia G. Electrodermal activity in antidepressant medicated and unmedicated depressive patients and in matched healthy subjects. Acta Psychiatr Scand 1987;76(6):684–692. doi: 10.1111/j.1600-0447.1987.tb02940.x

69. Abd-Alrazaq A, AlSaad R, Shuweihdi F, Ahmed A, Aziz S, Sheikh J. Systematic review and meta-analysis of performance of wearable artificial intelligence in detecting and predicting depression. Npj Digit Med Nature Publishing Group; 2023 May 5;6(1):1–16. doi: 10.1038/s41746-023-00828-5

70. Woll S, Birkenmaier D, Biri G, Nissen R, Lutz L, Schroth M, Ebner-Priemer UW, Giurgiu M. Applying AI in the Context of the Association Between Device-Based Assessment of Physical Activity and Mental Health: Systematic Review. JMIR MHealth UHealth 2025 Mar 6;13(1):e59660. doi: 10.2196/59660

71. Sempionatto JR, Lasalde-Ramírez JA, Mahato K, Wang J, Gao W. Wearable chemical sensors for biomarker discovery in the omics era. Nat Rev Chem Nature Publishing Group; 2022 Dec;6(12):899–915. doi: 10.1038/s41570-022-00439-w

72. Provenza NR, Sheth SA, Dastin-van Rijn EM, Mathura RK, Ding Y, Vogt GS, Avendano-Ortega M, Ramakrishnan N, Peled N, Gelin LFF, Xing D, Jeni LA, Ertugrul IO, Barrios-Anderson A, Matteson E, Wiese AD, Xu J, Viswanathan A, Harrison MT, Bijanki KR, Storch EA, Cohn JF, Goodman WK, Borton DA. Long-term ecological assessment of intracranial electrophysiology synchronized to behavioral markers in obsessive-compulsive disorder. Nat Med 2021 Dec;27(12):2154–2164. PMID:34887577

73. Provenza NR, Reddy S, Allam AK, Rajesh SV, Diab N, Reyes G, Caston RM, Katlowitz KA, Gandhi AD, Bechtold RA, Dang HQ, Najera RA, Giridharan N, Kabotyanski KE, Momin F, Hasen M, Banks GP, Mickey BJ, Kious BM, Shofty B, Hayden BY, Herron JA, Storch EA, Patel AB, Goodman WK, Sheth SA. Disruption of neural periodicity predicts clinical response after deep brain stimulation for obsessive-compulsive disorder. Nat Med Nature Publishing Group; 2024 Oct;30(10):3004–3014. doi: 10.1038/s41591-024-03125-0

74. Hurley ME, Sonig A, Herrington J, Storch EA, Lázaro-Muñoz G, Blumenthal-Barby J, Kostick-Quenet K. Ethical considerations for integrating multimodal computer perception and neurotechnology. Front Hum Neurosci Frontiers; 2024 Feb 16;18. doi: 10.3389/fnhum.2024.1332451

75. Kostick-Quenet KM, Herrington, John, and Storch EA. Personalized Roadmaps for Returning Results From Digital Phenotyping. Am J Bioeth Taylor & Francis; 2024 Feb 1;24(2):102–105. PMID:38295237

76. Martinez-Martin N, Insel TR, Dagum P, Greely HT, Cho MK. Data mining for health: staking out the ethical territory of digital phenotyping. Npj Digit Med Nature Publishing Group; 2018 Dec 19;1(1):1–5. doi: 10.1038/s41746-018-0075-8

77. Ortiz A, Halabi R, Alda M, Burgos A, DeShaw A, Gonzalez-Torres C, Husain MI, O’Donovan C, Tolend M, Hintze A, Mulsant BH. Day-to-day variability in sleep and activity predict the onset of a hypomanic episode in patients with bipolar disorder. J Affect Disord 2025 Jan 8; doi: 10.1016/j.jad.2025.01.026

78. Zhang Y, Pratap A, Folarin AA, Sun S, Cummins N, Matcham F, Vairavan S, Dineley J, Ranjan Y, Rashid Z, Conde P, Stewart C, White KM, Oetzmann C, Ivan A, Lamers F, Siddi S, Rambla CH, Simblett S, Nica R, Mohr DC, Myin-Germeys I, Wykes T, Haro JM, Penninx BWJH, Annas P, Narayan VA, Hotopf M, Dobson RJB. Long-term participant retention and engagement patterns in an app and wearable-based multinational remote digital depression study. Npj Digit Med Nature Publishing Group; 2023 Feb 17;6(1):1–13. doi: 10.1038/s41746-023-00749-3

79. Büscher R, Winkler T, Mocellin J, Homan S, Josifovski N, Ciharova M, van Breda W, Kwon S, Larsen ME, Torous J, Firth J, Sander LB. A systematic review on passive sensing for the prediction of suicidal thoughts and behaviors. Npj Ment Health Res Nature Publishing Group; 2024 Sept 23;3(1):1–10. doi: 10.1038/s44184-024-00089-4

80. Bladon S, Eisner E, Bucci S, Oluwatayo A, Martin GP, Sperrin M, Ainsworth J, Faulkner S. A systematic review of passive data for remote monitoring in psychosis and schizophrenia. Npj Digit Med Nature Publishing Group; 2025 Jan 27;8(1):1–13. doi: 10.1038/s41746-025-01451-2

81. Schyvens A-M, Peters B, Van Oost NC, Aerts J-M, Masci F, Neven A, Dirix H, Wets G, Ross V, Verbraecken J. A performance validation of six commercial wrist-worn wearable sleep-tracking devices for sleep stage scoring compared to polysomnography. Sleep Adv J Sleep Res Soc 2025 Apr;6(2):zpaf021. PMID:40303381

