## Supplementary Materials for "Digital Biomarkers for Passive Remote Monitoring of Bipolar Disorder: Systematic Review"

**Supplementary Methods:** Search terms

**Field restriction.** All terms are limited to the **Title/Abstract** field to prioritize topical relevance and reduce noise from full-text indexing. MeSH limits are used only for Humans and Adults.

**Concept blocks (combined as C1 AND C2 AND C3):**

- **C1: Bipolar disorder diagnosis terms.**
  Captures BD regardless of subtype and includes a core manic-state term:
  "bipolar disorder" [Title/Abstract] OR "bipolar depression" [Title/Abstract] OR "bipolar I disorder" [Title/Abstract] OR "bipolar II disorder" [Title/Abstract] OR "bipolar 1 disorder" [Title/Abstract] OR "bipolar 2 disorder" [Title/Abstract] OR mania [Title/Abstract]
- **C2: Behavioral/physiological signals.**
  Encompasses the feature domains used in remote/passive monitoring (mood/affect, sleep/circadian, activity/motor/mobility, socialization, speech/voice, vision/face, physiology incl. HR/HRV/EDA, light exposure, device usage):
  (mood [Title/Abstract] OR emotion [Title/Abstract] OR physiology [Title/Abstract] OR "heart rate" [Title/Abstract] OR electrophysiology [Title/Abstract] OR electrophysiological [Title/Abstract] OR behavior [Title/Abstract] OR activity [Title/Abstract] OR motor [Title/Abstract] OR sleep [Title/Abstract] OR energy [Title/Abstract] OR socialization [Title/Abstract] OR social [Title/Abstract] OR speech [Title/Abstract] OR movement [Title/Abstract] OR face [Title/Abstract] OR "facial expression" [Title/Abstract] OR location [Title/Abstract] OR geolocation [Title/Abstract] OR function [Title/Abstract] OR HRV [Title/Abstract] OR wellbeing [Title/Abstract] OR "device usage" [Title/Abstract] OR light [Title/Abstract])
- **C3: Monitoring/assessment intent.**
  Focuses results on measurement and biomarker aims rather than unrelated mentions:
  (tracking [Title/Abstract] OR monitoring [Title/Abstract] OR assessment [Title/Abstract] OR observation [Title/Abstract] OR detection [Title/Abstract] OR identification [Title/Abstract] OR marker [Title/Abstract] OR measuring [Title/Abstract])

**Filters.** Publication year, language, species, and age group were restricted as follows:

- AND ("2000" [Date - Publication] : "3000" [Date - Publication]) AND (English [Language]) AND (Humans [MeSH Terms]) AND (Adult [MeSH Terms])

PubMed example search:

| **Search terms** |
|---|
| ("bipolar disorder" [Title/Abstract] OR "bipolar depression" [Title/Abstract] OR "bipolar I disorder" [Title/Abstract] OR "bipolar II disorder" [Title/Abstract] OR "bipolar 1 disorder" [Title/Abstract] OR "bipolar 2 disorder" [Title/Abstract] OR mania [Title/Abstract]) |
| AND (mood [Title/Abstract] OR emotion [Title/Abstract] OR physiology [Title/Abstract] OR "heart rate" [Title/Abstract] OR electrophysiology [Title/Abstract] OR electrophysiological [Title/Abstract] OR behavior [Title/Abstract] OR activity [Title/Abstract] OR motor [Title/Abstract] OR sleep [Title/Abstract] OR energy [Title/Abstract] OR socialization [Title/Abstract] OR social [Title/Abstract] OR speech [Title/Abstract] OR movement [Title/Abstract] OR face [Title/Abstract] OR "facial expression" [Title/Abstract] OR location [Title/Abstract] OR geolocation [Title/Abstract] OR function [Title/Abstract] OR HRV [Title/Abstract] OR wellbeing [Title/Abstract] OR “device usage” [Title/Abstract] OR light) |
| AND (tracking [Title/Abstract] OR monitoring [Title/Abstract] OR assessment [Title/Abstract] OR observation [Title/Abstract] OR detection [Title/Abstract] OR identification [Title/Abstract] OR marker [Title/Abstract] OR measuring [Title/Abstract]) |
| AND ("2000" [Date - Publication] : "3000" [Date - Publication]) AND (English [Language]) AND (Humans [MeSH Terms]) AND (Adult [MeSH Terms]) |

**Supplementary Table 1:** Included Study Characteristics

| Author (Year) | Marker Category: Description | Sensing Methods | Statistical/ML Methods | Clinical Scales (Frequency) | Sample |
|---|---|---|---|---|---|
| Anmella (2024) | Electrodermal Activity: Patients with bipolar depression showed significantly reduced mean EDA and EDA peaks per minute, which increased to levels similar to euthymia or healthy controls after clinical remission | *Device*: Wearable (Empatica E4)<br>*Measurements*: Acceleration, skin temperature, blood volume pulse, heart rate, and electrodermal activity<br>*Data Characteristics*: 48hr recordings at acute episode and remission | ANOVA, paired t-tests | YMRS, HDRS (at 2 timepoints) | N = 49, inpatients |
| Faurholt-Jepsen (2014) | Geolocation: Change in smartphone cell tower ID is negatively correlated with depression severity. | *Device:* Smartphone (MONARCA app)<br>*Measurements:* Geolocation<br>*Data Characteristics:* Daily for 3 months | Mixed effect regression models | YMRS, HDRS (every 2 weeks) | N = 17, outpatients |
| Faurholt-Jepsen (2021) | Geolocation: Patients with BD during a depressive state were less mobile compared with a euthymic state. Patients with BD during an affective state had lower location entropy compared with a euthymic state | *Device:* Smartphone (MONARCA app)<br>*Measurements:* location data via GPS, WiFi, cell tower<br>*Data Characteristics:* mean of 101 days per patient | Mixed effect regression models | YMRS, HDRS (one time point) | N = 48, outpatients |
| Palmius (2017) | Geolocation: There is a strong link between geographic movements and depression in bipolar disorder, demonstrating an optimal mean absolute error rate when predicting clinical scale scores of 3.73, and a classification accuracy of 0.849. | *Device:* Smartphone (custom app on provided smartphone)<br>*Measurements:* anonymized geolocation data<br>*Data Characteristics:* daily for an average of 8.4 weeks in euthymia, 8.6 weeks in depressive patients | Linear regression models, a quadratic discriminant analysis classifier | QIDS (weekly) | N = 22, outpatient |
| Faurholt-Jepsen (2017) | Heart Rate: HRV is significantly higher in a manic state relative to depressive and euthymic states. | *Device:* Wearable (Actiheart)<br>*Measurements:* HRV<br>*Data Characteristics:* Minimum 3 continuous days during affective state | Mixed effect regression models | YMRS, HDRS (one time point) | N = 16, outpatients |
| Wazen (2018) | Heart Rate: Mania is associated with increased heart rate and decreased HRV compared to euthymia. | *Device:* Wearable *(*HRV monitor - Polar RS 800 CX frequencymeter)<br>*Measurements:* 20 minutes in supine position<br>*Data Characteristics:* performed in clinic during mania and at discharge | Wilcoxon's signed-rank test | BRMS (at admission and discharge) | n =19, inpatient, all male |
| Valenza (2014) | Heart Rate: Personalized monitoring models to recognize mood states with a total classification accuracy up to 95.81%. | *Device:* Wearable (custom system comprised of a t-shirt with integrated fabric electrodes and sensors)<br>*Measurements:* ECGs<br>*Data Characteristics:* worn in the afternoon till morning (18hr) 3-6 times | Markov chain models | HDRS, YMRS (2-3 times a month) | N = 8, inpatient |
| Valenza (2016) | Heart Rate: Personalized prediction accuracies in forecasting a mood state (euthymia/non-euthymia) at day t+1 were 69% on average, reaching values as high as 83.3%. | *Device:* Wearable (custom system comprised of a t-shirt with integrated fabric electrodes and sensors)<br>*Measurements:* ECGs<br>*Data Characteristics:* during the evening and night twice a week for 14 weeks | Support Vector Machines | HDRS, YMRS (at admission) | N = 14, inpatient |
| Nardelli (2017) | Heart Rate: There is a clear loss of complexity modulation of heartbeat dynamics during depressive and hypomanic states compared to euthymia. | *Device:* Wearable (Wearable system comprised of a comfortable t-shirt with integrated fabric electrodes and sensors)<br>*Measurements:* ECGs<br>*Data Characteristics:* daytime (8am-8pm), nighttime (8pm-7:30am) (2-3 times a month) | Multiscale entropy analysis | QIDS (2-3 times a month) | N = 8, inpatient |
| Corponi (2024) | Heart Rate: This paper establishes positive change in natural logarithm of the Root Mean Square of Successive RR interval Differences as a marker of symptom resolution regardless of episode polarity (95.175% probability of positive direction). | *Device*: Wearable (Empatica E4)<br>*Measurements*: Heart Rate Variability<br>*Data Characteristics*: 48 hour recordings at 4 acute timepoints | Bayesian model | YMRS, HDRS (up to four times: acute phase, clinical | N = 23, inpatients |

| | | | | | |
|---|---|---|---|---|---|
| | | | | response, remission, euthymia) | |
| Liu (2024) | Keyboard Usage: Higher smartphone keyboard backspace rates are associated with symptoms of mania and depression. Individuals were grouped into low, medium, and high backspace rate groups. The medium group had significantly higher ratings of depression, and both high and medium were significantly associated with higher ratings of mania | Device: Smartphone (BiAffect app on Android smartphones)<br>*Measurements:* backspace rates<br>*Data Characteristics:* used as primary keyboard for 4 weeks | Bayesian Mixture Model | HDRS, YMRS (week 2 and 4) | N = 101, outpatient |
| Stange (2018) | Keyboard Usage: Instability of typing speed predicted elevated future symptoms of depression, but not mania. Passive assessments of typing instability accounted for a similar amount of variance in future mood symptoms compared to actively reported EMA mood instability. | *Device:* Smartphone (BiAffect app on Android smartphones)<br>*Measurements:* keyboard strokes passively collected<br>*Data Characteristics:* prompted semi-randomly once a day over 2 weeks | Multilevel models (using MPlus), bootstrapped mediation analysis | HDRS, YMRS (weekly) | N = 18, outpatient |
| Zulueta (2018) | Keyboard Usage: Depression scores were positively correlated with accelerometer displacement, average interkey delay, session count, and autocorrect rate. For mania, accelerometer displacement is positively correlated and backspace rate is negatively correlated | Device: Smartphone (BiAffect app on Android smartphones)<br>Measurements: keyboard strokes passively collected<br>*Data Characteristics:* passively collected over 8 weeks | Mixed-effects regression model | HDRS, YMRS (weekly) | N = 9 (5 BD I, 4 BD II), outpatient |
| Cao (2017) | Keyboard Usage: This study uses a deep neural network trained on smartphone keyboard typing features to classify mild/severe depression (90% accuracy) and predict YMRS scores. | Device: Smartphone (BiAffect app on Android smartphones)<br>Measurements: keyboard strokes passively collected<br>*Data Characteristics:* passively collected over 8 weeks | A novel "DeepMood' deep learning architecture, utilizing Gated Recurrent Units (GRU) | HDRS, YMRS (weekly) | N = 12 (7 BD I, 5 BD II), outpatient |
| Esaki (2022) | Light Exposure: Bedroom light exposure at night is significantly associated with future manic episodes. Mania only. | *Device:* Portable Photometer (LX-28SD)<br>*Measurements:* light exposure<br>*Data Characteristics:* 7 consecutive days at baseline and follow-up | Cox proportional hazards model | YMRS (at baseline and at 2 year follow-up) | N = 157, outpatients |
| Esaki (2020) | Light Exposure: Bedroom light exposure at night is significantly associated with manic symptoms. Mania only. | *Device:* Portable Photometer (LX-28SD)<br>*Measurements:* light exposure<br>*Data Characteristics:* 7 consecutive days | t-test, Mann-Whitney U test | YMRS (one time point) | N = 184, outpatients |
| Esaki (2019) | Light Exposure: Higher daytime light intensity is associated with lower depression severity across (not within) patients. Depression only. | *Device:* Wearable (Actiwatch Spectrum Plus)<br>*Measurements:* light exposure<br>*Data Characteristics:* 7 consecutive days | t-test, Mann-Whitney U test | MADRS (one time point) | N = 181, inpatients |
| Esaki (2021a) | Light Exposure: Higher daytime light intensity is associated with lower likelihood of depression relapse across (not within) patients. Depression only. | *Device:* Wearable (Actiwatch Spectrum Plus)<br>*Measurements:* light exposure<br>*Data Characteristics:* 7 consecutive days | Cox proportional hazards model | MADRS (at baseline and at 12 month follow-up) | N = 198, outpatients |
| Ebner-Priemer (2020) | Physical Activity: Passively tracked activity metrics were statistically meaningfully associated with same-day manic and depressive psychopathology. | *Device:* Smartphone (movisensXS app)<br>*Measurements:* Incoming and outgoing phone calls and text messages, number of different call and text contacts, frequency and duration of times the display was on/off, rates of transmitted and received data, travel distances in kilometers, frequency and duration of different activity classes (in vehicle, on bicycle, walking, still, unknown, tilting) and the velocity of movement and number of steps<br>*Data Characteristics:* 12 month monitoring period, daily recordings | Multilevel structural equation monitoring | YMRS, BRMRS, MADRS (every 2 weeks) | N = 29, outpatients |
| Beiwinkel (2016) | Multi-Marker (Geolocation, Socialization): Smartphone-based measures (self-reported mood, social communication, and physical activity) were closely linked to both depressive and manic symptoms. Specifically, lower mood, fewer outgoing texts, | *Device:* Smartphone (Social Information Monitoring for Patients with Bipolar Affective Disorder app)<br>*Measurements:* movement activity: GPS, cell tower movement, accelerometry, and socialization: number and | Random-coefficient multilevel models | YMRS, HAMD (every 8 weeks) | N = 13, outpatients |

| | | | | | |
|---|---|---|---|---|---|
| | and reduced movement predicted higher or increasing depressive symptoms, while lower physical activity but higher social communication predicted higher or increasing manic symptoms. | duration of outgoing calls and the number of SMS sent per day<br>*Data Characteristics:* Daily data was collected for up to 12 months | | | |
| Faurholt-Jepsen (2016a) | Multi-Marker (Socialization, Geolocation): The more severe the depressive symptoms (1) the longer the smartphone's screen was "on"/day, (2) more received incoming calls/day, (3) fewer outgoing calls/day were made, (4) less answered incoming calls/day, (5) the patients moved less between cell towers IDs/day. Conversely, the more severe the manic symptoms (1) more outgoing text messages/day sent, (2) the phone calls/day were longer, (3) the fewer number of characters in incoming text messages/day, (4) the lower duration of outgoing calls/day, (5) the patients moved more between cell towers IDs/day. | *Device:* Smartphone (MONARCA app)<br>*Measurements:* the number of incoming text messages/day; the number of outgoing text messages/day; the duration of phone calls/day (collected with a maximum of 120 seconds/call); the amount of time the smartphone's screen was "on"/day; the number of times the smartphone's screen was turned "on"/day; and the number of cell tower ID changes/day<br>*Data Characteristics:* Daily for 12 weeks | Mixed effect regression models | YMRS, HDRS (every 2 weeks) | N = 29, outpatients |
| Anmella (2023) | Multi-Marker Classification (Physical Activity, Electrodermal Activity, Heart Rate): BiLSTM trained on wearable data at 3 acute timepoints acheived 61-70% accuracy at classifying severity of an acute affective episode. Most important features were accelleration, electrodermal activity, and HR. | *Device*: Wearable (Empatica E4)<br>*Measurements*: Acceleration, skin temperature, blood volume pulse, heart rate, and electrodermal activity<br>*Data Characteristics*: 48hr recordings at acute episode, response, and remission | Bidirectional Long-Short Term Memory neural network | YMRS, HDRS (at 3 timepoints) | N = 8, inpatients |
| Cote-Allard (2022) | Multi-Marker Classification (Physical Activity, Electrodermal Activity, Heart Rate): This work establishes a deep learning methodology that used wearable data to distinguish manic and euthymic states across (not within) patients. | *Device:* Wearable (Empatica E4)<br>*Measurements*: Accelerometry, Electrodermal Activity, PPG, HR<br>*Data Characteristics*: 24 hour recordings | Long–Short Term Memory | YMRS, MADRS (one time point) | N = 47, inpatients |
| Cho (2019) | Multi-Marker Classification (Physical Activity, Sleep, Heart Rate, Light Exposure): Features related to circadian rhythms were able to classify mood episodes in BDII and BDII with moderate accuracy. | *Device:* Wearable (Fitbit Charge), Smartphone (Android)<br>*Measurements:* 4 different categories related to circadian rhythms: light exposure, steps, sleep, and heart rate<br>*Data Characteristics:* 2 years of daily watch measurements | Random forest | YMRS, QIDS (every 12 weeks) | N = 37, outpatients |
| Ortiz (2025) | Multi-Marker (Physical Activity, Sleep): 12-hr variability in sleep (balanced accuracy = 0.87) and metabolic activity (balanced accuracy = 0.89) achieved high performance in detecting hypomanic episode onset, predicting the onset with a median delay of 3 and 2.5 days respectively. | *Device:* Wearable (Oura Health Oy, Generation 2)<br>*Measurements:* sleep stages in 5 minute intervals, metabolic activity in 5 minute intervals<br>*Data Characteristics:* collected continuously for a median of 495 days | Time-series oscillatory mode decomposition, Hilbert transform, and spectral derivative computation followed by peak detection to determine time-frequency variability | ASRM, PHQ-9 (weekly) | N = 50, outpatients |
| Lee (2023) | Multi-Marker Classification (Sleep, Physical Activity, Heart Rate, Light Exposure): This study aims to use circadian rhythm disruptions measured through aggregated wearable data to classify upcoming depressive/manic episodes over the next 3 days. The prediction accuracies for impending major depressive episodes was 90.1% (AUC of 0.937), manic episodes was 92.6% (AUC of 0.957), and hypomanic episodes was 93% (AUC of 0.963) for the next 3 days | *Device:* Wearable (Fitbit Charge), Smartphone (Android)<br>*Measurements:* sleep, steps, heart rate, ambient light<br>*Data Characteristics:* continuously for a minimum of 30 days, average of 279.7 days | Random Forest | YMRS, MADRS (every 3 months) | N = 175, (78 BD I, 97 BD II), inpatient |
| Lim (2024) | Multi-Marker Classification (Sleep): Features related to sleep and circadan rhythms enabled accurate next-day predictions for depressive, manic, and hypomanic episodes (AUCs: 0.80, 0.98, 0.95). Daily circadian phase shifts were the most significant predictors: delays linked to depressive episodes, advances to manic episodes. | *Device:* Wearable (Fitbit Charge)<br>*Measurements:* sleep derived 36 sleep and circadian rhythm features<br>*Data Characteristics:* minimum 30 days per patient, average of 267 days | XGBoost | YMRS, MADRS (every 3 months) | N = 111, outpatient |

| | | | | | |
|---|---|---|---|---|---|
| Lipschitz (2024) | Multi-Marker Classification (Sleep, Physical Activity, Heart Rate): Aggregated wearable data was able to classify depression (AUC=0.86) and mania (AUC=0.85) | *Device:* Wearable (Fitbit Inspire)<br>*Measurements:* 17 features related to sleep, activity, and HR<br>*Data Characteristics:* continuously for 9 months | Binary Mixed Model forest | ASRM, PHQ-8 (biweekly) | N = 54, outpatient |
| Esaki (2021b) | Physical Activity: A later timing of circadian activity rhythm (M10 onset time) was significantly associated with future depressive episode relapses. | *Device:* Wearable (Actiwatch Spectrum Plus)<br>*Measurements:* Accelerometry<br>*Data Characteristics:* 7 consecutive days at baseline and follow-up | Cox proportional hazards model | YMRS, MADRS (at baseline and at 12 month follow-up) | N = 189, outpatients |
| Gershon (2016) | Physical Activity: Depressive days are distinguished from other mood states by an overall lower activity level, and a pattern of later activity onset, a midday elevation of activity, and low evening activity. No distinct within-person activity patterns were found for hypomanic/manic days. | *Device:* Wearable (Actiwatch)<br>*Measurements:* accelerometry<br>*Data Characteristics:* 2 consecutive months | Functional principal components analysis | YMRS, IDS-C (every month) | N = 37, outpatients |
| Jakobsen (2022) | Physical Activity: Motor activity of mania is characterized by altered complexity and variability when compared within-subject to euthymia. | *Device:* Wearable (Empatica E4)<br>*Measurements:* accelerometry<br>*Data Characteristics:* 24 hour recordings at admission and remission | Paired-Samples T-tests, Wilcoxon Signed Rank Test | YMRS, MADRS (2 times) | N = 16, inpatients |
| Faurholt-Jepsen (2016b) | Physical Activity: There was a significant correlation between the severity of manic symptoms and energy expenditure and trunk acceleration, respectively (across patients). | *Device:* Wearable (Actiheart, mounted on thorax)<br>*Measurements:* Psychomotor activity (movement/acceleration (m/s2) and heart rate (bpm)<br>*Data Characteristics:* Minimum 3 continuous days during affective state | Mixed effect regression models | YMRS, HDRS (one time point) | N = 19, outpatients |
| Esaki (2023) | Sleep: Variability in total sleep time was significantly associated with an increase in the mood episode relapses, and participants with higher variability in total sleep time had a significantly shorter mean estimated time to mood episode relapse. | *Device:* Wearable (Actiwatch Spectrum Plus)<br>*Measurements:* Accelerometry<br>*Data Characteristics:* 7 consecutive days at baseline and follow-up | Cox proportional hazards model | YMRS, MADRS (at baseline and at 2 year follow-up) | N = 193, outpatients |
| Esaki (2024) | Sleep: There is a significant association between daytime napping and depressive symptoms in patients with bipolar disorder. Specifically, in a multivariable logistic regression analysis, as the number of nap days, number of naps per day, and nap duration increased, the odds ratio (OR) for depressed state significantly increased. | *Device:* Wearable (Actiwatch Spectrum Plus)<br>*Measurements:* Accelerometry<br>*Data Characteristics:* 7 consecutive days at baseline | Cox proportional hazards model | YMRS, MADRS (at baseline) | N = 204, outpatients |
| Dominiak (2022) | Socialization: Smartphone socialization data collected in the 7 days before and 2 days after a clinical visit shows significant differences in depressive, euthymic, and mixed/manic states. | *Device:* Smartphone (BDmon app)<br>*Measurements*: Calls and text message statistics, including incoming answered and missed calls, and outgoing calls and text messages<br>*Data Characteristics*: Data was collected for the 7 days before and 2 days after a clinical evaluation, with a minimum of one evaluation per participant | Linear mixed-effects model | YMRS, HDRS (every 3 months, or after a mood change was suspected after a fortnightly phone call intervention) | N = 51, outpatients |
| Grünerbl (2015) | Multi-Marker Classification (Socialization, Geolocation, Physical Activity, Speech): The aim of the study was to identify recognition of states and changes. Using socialization features, we gain recognition accuracies of 76% by fusing all sensor modalities and state change detection precision and recall of over 97%. | *Device:* Smartphone (Android smartphone running custom logging application)<br>*Measurements:* number of phone calls, total length, average length, number of unique numbers<br>*Data Characteristics:* 12 weeks continously, over 800 days total | Naive Bayes classifier | HAMD, YMRS (every 3 weeks) | N = 10, inpatient |
| Faurholt-Jepsen (2015) | Socialization: There are significant correlations between smartphone socialization metrics and depressive and manic symptoms. | *Device:* Smartphone (MONARCA app)<br>*Measurements:* number of incoming and outgoing phone calls per day, the duration of incoming and outgoing phone calls per day, and the number of outgoing text messages per day | Mixed effect regression models | YMRS, HDRS (every 2 weeks) | N = 61, outpatients |

| | | | | | |
|---|---|---|---|---|---|
| | | *Data Characteristics:* Daily for 6 months | | | |
| Ji (2024) | Speech: Detected depressive and manic states from real-world journal voice recordings with an overall accuracy of 86.1% for detecting depression and 90.3% for mania. | *Device:* Smartphone (MoodMirror app)<br>*Measurements:* audio recordings<br>*Data Characteristics:* prompted once a day | Chinese-speech-pretrained Gate Recurrent Unit model | QIDS, YMRS (on app daily) | N = 93, inpatient |
| Faurholt-Jepsen (2016c) | Speech: Affective states were classified using voice features extracted during everyday life phone calls. Voice features were found to be more accurate, sensitive and specific in the classification of manic or mixed states (AUC = 0.89) compared to depressive states (AUC = 0.78). | *Device:* Smartphone (MONARCA app, openSMILE toolkit)<br>*Measurements:* voice features extracted from natural phone calls<br>*Data Characteristics:* Daily for 12 weeks | Random forest | YMRS, HDRS (every 2 weeks) | N = 28, outpatients |
| Karam (2014) | Speech: Classified mania (AUC=0.81) and depression (AUC=0.67) solely using speech colleced during clinical evaluations. | *Device:* Smartphone (custom app)<br>*Measurements:* records outgoing speech at 8KhZ<br>*Data Characteristics:* 6-31 weeks of weekly calls | Support Vector Machines | HDRS, YMRS (weekly over phone) | N = 6, inpatient |
| Zhang (2018) | Speech: Established characteristic speech signals that differed between manic and euthymic groups. Specifically, the fourth formant and Linear Prediction Coefficient were significantly differed when patients progressed from manic to remission state. | *Device:* Smartphone (custom app developed by team on provided Android mobile phones)<br>*Measurements:* audio recording outgoing calls in a quiet indoor room<br>*Data Characteristics:* 25-30 min clinical phone conversations, two times | Two-sided t-test, chi-square tests | BRMS (after 2 audio collection sessions) | N = 30, outpatient |
| Gideon (2016) | Speech: Speech features can be used to classify manic (AUC = 0.72) and depressive (AUC = 0.75) episodes during clinical interviews | *Device:* Smartphone (PRIORI app)<br>*Measurements:* Speech<br>*Data Characteristics:* 6-12 months | Support Vector Machines | YMRS, HAMD (weekly) | N = 37, outpatients |
| Pan (2018) | Speech: Spontaneous conversations with clinicians were used to classify manic vs euthymic states, where Support Vector Machines were superior for classification within individuals (88.56% accuracy) and Gaussian Mixture Models were superior for classification across individuals (72.27% accuracy). | *Device:* Smartphone (preloaded Samsung GALAXY Mega 6.3, calls recorded over Cloud network)<br>*Measurements:* clinican would have free open conversation which was recorded and transferred to a cloud database<br>*Data Characteristics:* recorded for 10-25 minutes twice in each mood state (manic and euthymic) in the morning in consecutive 1–2 days | Support Vector Machines, Gaussian Mixture Models | BRMS (at admission) | N = 21, inpatient |
| Kaczmarek-Majer (2024) | Speech: The study showed that prosodic, spectral, and voice quality parameters are all valid<br>markers in assessing the severity of manic and depressive symptoms. The greater the severity of mania in males, the louder and higher the tone of voice. For females, the greater the severity of mania, the quieter and lower the tone of voice. | *Device:* Smartphone (BDmon app)<br>*Measurements:* speech during all phone calls<br>*Data Characteristics:* phone calls every fortnight, used on average of 208 days | Generalized mixed-effect model | HDRS, YMRS (every 3 months, and additionally when a mood change was suspected during fortnightly phone calls) | N = 51, mix of inpatient and outpatient |
| Siegel (2024) | Speech: This study demonstrates that there are slower speech latencies in bipolar depression, with speech latencies significantly correlating with MADRS scores (r=0.3). | *Device:* Microphone<br>*Measurements:* audio recordings during clinical interviews<br>*Data Characteristics:* recorded on weeks 1, 2, 3, 4, and 6, (interviews averaged 21.63 min) | Recursive Decision Tree, Multilevel models | MADRS (screening, baseline, weeks 1, 2, 3, 4, and 6) | N = 274, inpatient |

**Supplementary Table 2:** PROBAST Assessment

| Author (Year) | Participants | Predictors | Outcome | Analysis | Overall |
|---|---|---|---|---|---|
| Valenza (2016) | Low | Low | Low | High | High |
| Ortiz (2025) | Low | Low | Low | High | High |
| Lim (2024) | Low | Low | Low | High | High |
| Palmius (2017) | Low | Low | Low | High | High |
| Anmella (2023) | Low | Low | Low | High | High |
| Cote-Allard (2022) | Low | Low | Low | High | High |
| Cho (2019) | Low | Low | Low | High | High |
| Lipschitz (2024) | Low | Low | Low | High | High |
| Ji (2024) | Low | Low | Low | High | High |
| Faurholt-Jepsen (2016c) | Low | Low | Low | High | High |
| Karam (2014) | Low | Low | Low | High | High |
| Gideon (2016) | Low | Low | Low | High | High |
| Pan (2018) | Low | Low | Low | High | High |
| Cao (2017) | Low | Low | Low | High | High |
| Grünerbl (2015) | Low | Low | Low | High | High |

**Supplementary Table 3:** Newcastle-Ottawa Assessment

| Author (Year) | Study Type | Selection | Comparability | Exposure (cc) | Outcome (co) |
|---|---|---|---|---|---|
| Esaki (2021a) | Cohort | Representative, Exposure (2) | 2 | | Assessment, Follow-Up, Adequately (3) |
| Gershon (2016) | Cohort | Representative, Control, Exposure, Outcome (4) | 2 | | Assessment, Follow-up (2) |
| Jakobsen (2022) | Cohort | Representative, Exposure, Outcome (3) | 2 | | Assessment (1) |
| Anmella (2024) | Cohort | Representative, Exposure, Outcome (3) | 2 | | Assessment (1) |
| Faurholt-Jepsen (2014) | Cohort | Representative, Exposure, Outcome (3) | 0 | | Assessment, Follow-Up, Adequately (3) |
| Faurholt-Jepsen (2021) | Cohort | Representative, Control, Exposure, Outcome (4) | 0 | | Assessment (1) |
| Faurholt-Jepsen (2017) | Cohort | Representative, Exposure, Outcome (3) | 2 | | Assessment, Follow-up (2) |
| Nardelli (2017) | Cohort | Representative, Exposure (2) | 0 | | Assessment, Follow-Up, Adequately (3) |
| Corponi (2024) | Cohort | Representative, Exposure, Outcome (3) | 2 | | Assessment, Follow-Up, Adequately (3) |
| Liu (2024) | Cohort | Representative, Exposure (2) | 2 | | Assessment, Follow-Up, Adequately (3) |
| Stange (2018) | Cohort | Representative, Exposure (2) | 2 | | Assessment, Follow-up (2) |
| Zulueta (2018) | Cohort | Representative, Exposure (2) | 2 | | Assessment, Follow-Up, Adequately (3) |
| Esaki (2022) | Cohort | Representative, Exposure, Outcome (3) | 2 | | Assessment, Follow-Up, Adequately (3) |
| Esaki (2020) | Cohort | Representative, Exposure, Outcome (3) | 2 | | Assessment, Follow-Up, Adequately (3) |
| Esaki (2019) | Cohort | Representative, Exposure, Outcome (3) | 2 | | Assessment, Follow-Up, Adequately (3) |
| Esaki (2021b) | Cohort | Representative, Exposure, Outcome (3) | 2 | | Assessment, Follow-Up, Adequately (3) |
| Faurholt-Jepsen (2016a) | Cohort | Representative, Exposure, Outcome (3) | 2 | | Assessment, Follow-Up, Adequately (3) |
| Beiwinkel (2016) | Cohort | Representative, Exposure, Outcome (3) | 2 | | Assessment, Follow-Up, Adequately (3) |
| Faurholt-Jepsen (2016b) | Cohort | Representative, Exposure, Outcome (3) | 2 | | Assessment, Follow-up (2) |
| Esaki (2023) | Cohort | Representative, Exposure, Outcome (3) | 2 | | Assessment, Follow-Up, Adequately (3) |
| Dominiak (2022) | Cohort | Representative, Exposure, Outcome (3) | 2 | | Assessment, Follow-Up, Adequately (3) |
| Faurholt-Jepsen (2015b) | Cohort | Representative, Exposure, Outcome (3) | 2 | | Assessment, Follow-Up, Adequately (3) |
| Zhang (2018) | Cohort | Representative, Exposure, Outcome (3) | 2 | | Assessment, Follow-Up, Adequately (3) |
| Kaczmarek-Majer (2024) | Cohort | Representative, Exposure, Outcome (3) | 2 | | Assessment, Follow-Up, Adequately (3) |
| Siegel (2024) | Cohort | Representative, Exposure, Outcome (3) | 2 | | Assessment, Follow-Up, Adequately (3) |
| Wazen (2018) | Cohort | Exposure, Outcome (2) | 1 | | Assessment, Follow-Up, Adequately (3) |
| Valenza (2014) | Cohort | Representative, Exposure, Outcome (3) | 2 | | Assessment, Follow-up (2) |
| Ebner-Priemer (2020) | Cohort | Representative, Exposure, Outcome (3) | 2 | | Assessment, Follow-Up, Adequately (3) |
| Esaki (2024) | Cohort | Representative, Exposure, Outcome (3) | 2 | | Assessment, Follow-Up, Adequately (3) |

**Supplementary Table 4:** PRISMA Checklist

| Section and Topic | Item # | Checklist item | Page Number(s) or location where item is reported |
|---|---|---|---|
| **TITLE** | | | |
| Title | 1 | Identify the report as a systematic review. | 1 |
| **ABSTRACT** | | | |
| Abstract | 2 | See the PRISMA 2020 for Abstracts checklist. | 2 |
| **INTRODUCTION** | | | |
| Rationale | 3 | Describe the rationale for the review in the context of existing knowledge. | 3-5 |
| Objectives | 4 | Provide an explicit statement of the objective(s) or question(s) the review addresses. | 3-5 |
| **METHODS** | | | |
| Eligibility criteria | 5 | Specify the inclusion and exclusion criteria for the review and how studies were grouped for the syntheses. | 31 |
| Information sources | 6 | Specify all databases, registers, websites, organisations, reference lists and other sources searched or consulted to identify studies. Specify the date when each source was last searched or consulted. | 31 |
| Search strategy | 7 | Present the full search strategies for all databases, registers and websites, including any filters and limits used. | Supplementary Table 2 |
| Selection process | 8 | Specify the methods used to decide whether a study met the inclusion criteria of the review, including how many reviewers screened each record and each report retrieved, whether they worked independently, and if applicable, details of automation tools used in the process. | 32 |
| Data collection process | 9 | Specify the methods used to collect data from reports, including how many reviewers collected data from each report, whether they worked independently, any processes for obtaining or confirming data from study investigators, and if applicable, details of automation tools used in the process. | 32 |
| Data items | 10a | List and define all outcomes for which data were sought. Specify whether all results that were compatible with each outcome domain in each study were sought (e.g. for all measures, time points, analyses), and if not, the methods used to decide which results to collect. | 32-33 |
| | 10b | List and define all other variables for which data were sought (e.g. participant and intervention characteristics, funding sources). Describe any assumptions made about any missing or unclear information. | 32-33 |
| Study risk of bias assessment | 11 | Specify the methods used to assess risk of bias in the included studies, including details of the tool(s) used, how many reviewers assessed each study and whether they worked independently, and if applicable, details of automation tools used in the process. | 32-33 |
| Effect measures | 12 | Specify for each outcome the effect measure(s) (e.g. risk ratio, mean difference) used in the synthesis or presentation of results. | 32-33 |
| Synthesis methods | 13a | Describe the processes used to decide which studies were eligible for each synthesis (e.g. tabulating the study intervention characteristics and comparing against the planned groups for each synthesis (item #5)). | 32-33 |
| | 13b | Describe any methods required to prepare the data for presentation or synthesis, such as handling of missing summary statistics, or data conversions. | 32-33 |
| | 13c | Describe any methods used to tabulate or visually display results of individual studies and syntheses. | 32-33 |
| | 13d | Describe any methods used to synthesize results and provide a rationale for the choice(s). If meta-analysis was performed, describe the model(s), method(s) to identify the presence and extent of statistical heterogeneity, and software package(s) used. | 32-33 |
| | 13e | Describe any methods used to explore possible causes of heterogeneity among study results (e.g. subgroup analysis, meta-regression). | 32-33 |
| | 13f | Describe any sensitivity analyses conducted to assess robustness of the synthesized results. | 32-33 |
| Reporting bias assessment | 14 | Describe any methods used to assess risk of bias due to missing results in a synthesis (arising from reporting biases). | 32-33 |
| Certainty assessment | 15 | Describe any methods used to assess certainty (or confidence) in the body of evidence for an outcome. | 32-33 |

| Section and Topic | Item # | Checklist item | Page Number(s) or location where item is reported |
|---|---|---|---|
| **RESULTS** | | | |
| Study selection | 16a | Describe the results of the search and selection process, from the number of records identified in the search to the number of studies included in the review, ideally using a flow diagram. | 6 |
| | 16b | Cite studies that might appear to meet the inclusion criteria, but which were excluded, and explain why they were excluded. | n/a |
| Study characteristics | 17 | Cite each included study and present its characteristics. | 6-20 |
| Risk of bias in studies | 18 | Present assessments of risk of bias for each included study. | 20-21 |
| Results of individual studies | 19 | For all outcomes, present, for each study: (a) summary statistics for each group (where appropriate) and (b) an effect estimate and its precision (e.g. confidence/credible interval), ideally using structured tables or plots. | 6-20 |
| Results of syntheses | 20a | For each synthesis, briefly summarise the characteristics and risk of bias among contributing studies. | 6-20 |
| | 20b | Present results of all statistical syntheses conducted. If meta-analysis was done, present for each the summary estimate and its precision (e.g. confidence/credible interval) and measures of statistical heterogeneity. If comparing groups, describe the direction of the effect. | n/a |
| | 20c | Present results of all investigations of possible causes of heterogeneity among study results. | 6-20 |
| | 20d | Present results of all sensitivity analyses conducted to assess the robustness of the synthesized results. | n/a |
| Reporting biases | 21 | Present assessments of risk of bias due to missing results (arising from reporting biases) for each synthesis assessed. | n/a |
| Certainty of evidence | 22 | Present assessments of certainty (or confidence) in the body of evidence for each outcome assessed. | n/a |
| **DISCUSSION** | | | |
| Discussion | 23a | Provide a general interpretation of the results in the context of other evidence. | 22-30 |
| | 23b | Discuss any limitations of the evidence included in the review. | 22-30 |
| | 23c | Discuss any limitations of the review processes used. | 22-30 |
| | 23d | Discuss implications of the results for practice, policy, and future research. | 22-30 |
| **OTHER INFORMATION** | | | |
| Registration and protocol | 24a | Provide registration information for the review, including register name and registration number, or state that the review was not registered. | 6 |
| | 24b | Indicate where the review protocol can be accessed, or state that a protocol was not prepared. | 6 |
| | 24c | Describe and explain any amendments to information provided at registration or in the protocol. | n/a |
| Support | 25 | Describe sources of financial or non-financial support for the review, and the role of the funders or sponsors in the review. | 34 |
| Competing interests | 26 | Declare any competing interests of review authors. | 34 |
| Availability of data, code and other materials | 27 | Report which of the following are publicly available and where they can be found: template data collection forms; data extracted from included studies; data used for all analyses; analytic code; any other materials used in the review. | 34 |

*From:* Page MJ, McKenzie JE, Bossuyt PM, Boutron I, Hoffmann TC, Mulrow CD, et al. The PRISMA 2020 statement: an updated guideline for reporting systematic reviews. BMJ 2021;372:n71. doi: 10.1136/bmj.n71. This work is licensed under CC BY 4.0. To view a copy of this license, visit https://creativecommons.org/licenses/by/4.0/
